# Comprehensive Assessment of Sleep Duration, Insomnia and Brain Structure within the UK Biobank Cohort

**DOI:** 10.1101/2023.06.19.23291496

**Authors:** Aleks Stolicyn, Laura M. Lyall, Donald M. Lyall, Nikolaj Høier, Mark J. Adams, Xueyi Shen, James H. Cole, Andrew M. McIntosh, Heather C. Whalley, Daniel J. Smith

**Author notes:** Corresponding author: Prof Daniel Smith Division of Psychiatry, Centre for Clinical Brain Sciences, University of Edinburgh Kennedy Tower, Royal Edinburgh Hospital, Morningside Park, Edinburgh EH10 5HF, UK.

## Abstract

**Study Objectives:** To assess for associations between sleeping more than or less than recommended by the National Sleep Foundation (NSF), and self-reported insomnia, with brain structure.

**Methods:** Data from the UK Biobank cohort were analysed (*N* between 9K and 32K, dependent on availability, aged 44 to 82 years). Sleep measures included self-reported adherence to NSF guidelines on sleep duration (sleeping between 7 and 9 hours per night), and self-reported difficulty falling or staying asleep (insomnia). Brain structural measures included global and regional cortical or subcortical morphometry (thickness, surface area, volume), global and tract-related white matter microstructure, brain age gap (difference between chronological age and age estimated from brain scan), and total volume of white matter lesions.

**Results:** Longer-than-recommended sleep duration was associated with lower overall grey and white matter volumes, lower global and regional cortical thickness and volume measures, higher brain age gap, higher volume of white matter lesions, higher mean diffusivity globally and in thalamic and association fibers, and lower volume of the hippocampus. Shorter-than-recommended sleep duration was related to higher global and cerebellar white matter volumes, lower global and regional cortical surface areas, and lower fractional anisotropy in projection fibers. Self-reported insomnia was associated with higher global grey and white matter volumes, and with higher volumes of the amygdala, hippocampus and putamen.

**Conclusions:** Sleeping longer than recommended by the NSF is associated with a wide range of differences in brain structure, potentially indicative of poorer brain health. Sleeping less than recommended is distinctly associated with lower cortical surface areas. Future studies should assess the potential mechanisms of these differences and investigate long sleep duration as a putative marker of brain health.

## INTRODUCTION

Sleep is fundamentally important for physical and mental health. Sleep deprivation or insufficient sleep is associated with increased blood pressure,^1^ weight gain,^2,3^ increased risk of diabetes and heart disease,^4–6^ and immune system dysfunction.^7^ For mental health, problems with sleep (both hypersomnia or insomnia) are a feature of many psychiatric disorders, particularly dementia, schizophrenia, bipolar disorder, major depression and anxiety.^8–11^

Sleep has a multitude of functions, including the restoration of brain energy reserves,^12,13^ clearance of metabolic by-products of wakefulness,^14–16^ and the maintenance of attention and memory functions.^17–20^

Several studies have investigated relationships between sleep and brain structure. Lower sleep quality and the presence of sleep problems was associated with reduced hippocampal volumes over the lifespan in the Lifebrain consortium (*N* = 1299),^21^ while longer sleep duration was related to lower overall cerebral brain volumes in the Framingham Heart Study (*N* = 2060).^22^ For a sub-sample of the Human Connectome Project (*N* = 974), shorter sleep duration and poorer sleep quality were associated with lower intracortical myelin in parts of the cingulate, middle temporal and orbitofrontal cortices.^23^ In several smaller-scale studies poor sleep quality and non-optimal sleep duration have been associated with reduced grey matter volumes in frontal, temporal and parietal regions, as well as reduced white-matter integrity.^24,25^ Taken together, these findings indicate that sleep duration is non-linearly related to brain structure, with optimal sleep times associated with greater brain regional volumes.^22–24,26^ Poorer sleep quality, on the other hand, is likely related to lower cortical and subcortical volumes.^21,24^

Very few studies to date have looked at how sleep may be related to complex measures of brain structure which incorporate multiple imaging-derived phenotypes, such as brain-predicted age.^27^ For example, Ramduny et al. found an association of poor sleep quality with accelerated brain ageing in a small sample of older participants (*N* = 50),^28^ while Chu et al. found an association of acute sleep deprivation with changes in brain morphology reminiscent of advanced brain ageing (*N* = 134 young adults).^29^ These results have not yet been replicated in larger samples.

For insomnia, a recent ENIGMA consortium study found depression-specific insomnia severity to be associated with widespread decreases in cortical surface areas (*N* = 1053).^30^ In a Spanish study of risk for Alzheimer’s (ALFA, *N* > 330), diagnosis of insomnia was related to lower grey matter volume in the thalamus, cingulate and temporal cortices.^31^ The same study found associations with higher volume in the left caudate and widespread differences in white matter microstructure. These findings indicate that insomnia-related differences in the brain may depend on insomnia severity, as well as the presence of co-morbid conditions such as depression or Alzheimer’s disease.

Overall, studies of sleep and brain structure have so far been somewhat inconsistent, and brain measures of both grey and white matter structure have only rarely been studied within the same sample.^31^ We aimed to address these limitations by testing associations of sleep duration and insomnia with a comprehensive range of brain structural measures in the large UK Biobank cohort.

## METHODS

### Study Overview

We performed cross-sectional analyses to investigate associations of measures of grey matter structure and white matter microstructure with self-reported sleep variables in the UK Biobank.^32^ Drawing on previous findings, we hypothesised that suboptimal sleep duration (either too long or too short) and insomnia would be associated with reductions in grey matter volumes and differences in white matter microstructure. We aimed to better characterise these differences by leveraging the wide range of brain measures and the large sample sizes available within UK Biobank.

### Participant Sample

All data analysed in the current study were collected as part of the first imaging assessment conducted between April 2014 and March 2020. Participants were excluded from any analysis where they had missing sleep-related, covariate or brain imaging data, or where they had outlier values in any brain measures. Exclusions were performed separately for each group of outcome (brain imaging) variables to maximise sample sizes. No further exclusion criteria were applied. All primary analyses had between 9K and 32K participants. Table 1 presents characteristics of the sample used in the analyses of global grey and white matter volumes in relation to sleep guideline adherence (*N* = 32,119). Supplementary section S1.6 and the accompanying CSV tables provide further details of participant exclusions and the summary sample demographic characteristics for all conducted analyses. UK Biobank received ethical approval from the North West Multi-centre Research Ethics Committee in the UK (reference 11/NW/0382), and the current study received approval from the UKB Access Committee (application #4844). All participants gave written informed consent.

**Table 1.** Characteristics of the sample in the analyses of global grey and white matter volumes in relation to NSF guideline adherence with the main statistical model (please see supplementary section S1.6 for characteristics of the samples in all other analyses)

| Sample characteristic | Participants sleeping as NSF recommended (7-9 hours per day) | Participants sleeping less than NSF recommended (< 7 hours per day) | Participants sleeping more than NSF recommended (> 9 hours per day) |
| --- | --- | --- | --- |
| Sample size (male / female) | 24057<br>(11557 / 12500) | 7682<br>(3338 / 4344) | 380<br>(182 / 198) |
| Age mean (SD) | 63.65 (7.49) | 62.81 (7.62) * | 64.85 (7.53) * |
| Ethnic minority count (proportion) | 1931 (8.03%) | 742 (9.66%) * | 32 (8.42%) |
| Townsend index mean (SD) | -2.01 (2.65) | -1.77 (2.77) * | -1.53 (2.95) * |
| BMI mean (SD) | 26.35 (4.26) | 26.88 (4.66) * | 28.37 (5.49) * |
| Vascular / heart problems medicated count (proportion) | 7892 (32.81%) | 2519 (32.79%) | 178 (46.84%) * |
| Diabetic count (proportion) | 1172 (4.87%) | 433 (5.64%) * | 46 (12.11%) * |
| Smoking count (proportion) | 8804 (36.60%) | 2873 (37.40%) | 153 (40.26%) |
| Alcohol consumer count (proportion) | 17479 (72.66%) | 5299 (68.98%) * | 248 (65.26%) * |
| Lifetime depression count (proportion) | 5508 (22.90%) | 1980 (25.77%) * | 150 (39.47%) * |

**Note:** Significant differences in groups when compared to participants sleeping the NSF recommended time (less than 7 hours per day / more than 9 hours compared to 7-9 hours) are marked with asterisks. Characteristics were compared between groups with two-sample t-tests (for continuous variables) or with chi-squared tests (for participant proportions). Age is measured in years.

### Sleep Measures

Drawing on previous work,^21–26,30,31^ we focused our main analyses on three binary sleep-related variables: 1) sleeping longer than recommended by the National Sleep Foundation (NSF) for adults^33^ (> 9 hours per day, as compared to 7-9 hours); 2) sleeping less than recommended by the NSF (< 7 hours per day, as compared to 7-9 hours); and 3) insomnia/sleeplessness defined as usual trouble falling or staying asleep at night. As supplementary analyses, we also tested associations of sleep duration, longer compared to shorter than recommended sleep, late chronotype, and ‘ease of getting up’ (supplementary section S2.2). All variables were based on self-report data from the UK Biobank touchscreen questionnaire, collected as part of the first imaging assessment.^34^ Supplementary section S1.1 provides detailed definitions of sleep-related variables.

### Brain Structure Measures

We tested a set of global brain measures, a set of regional grey matter morphometric measures, and a set of white matter microstructure measures for individual tracts.

Global grey matter measures included the overall grey matter volume (head-size normalised), and global cortical thickness, surface area and cortical volume measures. Global white matter measures included overall white matter volume (head-size normalised), overall volume of white matter hyperintensities (lesions), and five PCA-derived measures of global white matter microstructure: fractional anisotropy (FA), mean diffusivity (MD), intracellular volume fraction (ICVF), orientation dispersion (OD) and isotropic volume fraction (ISOVF). Finally, a brain age gap estimate was derived for a subset of participants as the difference between age estimated from T1-weighted MRI scan (brain-predicted age) and actual chronological age (additionally residualised for chronological age). Brain-predicted age measures were obtained with brainageR toolkit version 2.1, through application of a predictive model based on Gaussian processes regression to normalised voxel-wise measures of grey matter, white matter and cerebrospinal fluid^27^ (supplementary section S1.2.1).

Individual-region grey matter morphometric data consisted of cortical thickness, surface area and volume measures for 5 bilateral lobes, 33 bilateral individual regions defined according to the Desikan-Killiany atlas, and volumes of 10 bilateral subcortical structures. White matter microstructure data consisted of FA and MD measures for three fiber bundles (association, projection and thalamic), and for 15 individual white matter tracts (12 bilateral and 3 unilateral). Supplementary analyses tested associations of fiber and tract-related ICVF, OD and ISOVF measures (supplementary section S2.1). Supplementary section S1.2 provides further details of the analysed brain measures.

### Statistical Models and Covariates

Analyses of unilateral (singular) brain measures were performed using generalized linear regression models (GLM). Analyses of bilateral measures were performed with linear mixed effects models (LME), with data for left and right hemispheres treated as repeated measures. Functions ‘fitglm’ and ‘fitlme’ in MATLAB R2018a (http://www.mathworks.com/products/matlab/, Mathworks Inc, RRID:SCR_001622) were used respectively for GLM and LME analyses. Brain measures were entered as outcome variables and sleep measures were entered as exposure variables. All data were standardised prior to analyses and all effect sizes are reported as standardised beta coefficients.

All statistical analyses included correction for age, sex, ethnic background, Townsend deprivation index, a set of health-related covariates, and three head position coordinates in the scanner. Analyses of cortical thickness, surface area and volume measures included an additional covariate of intracranial volume (ICV), standardised across the entire available sample, to correct for overall head size. Due to predominance of white British participants in the UK Biobank, ethnic background was defined as a binary variable indicating whether a participant was white British or not.^35^ Townsend index was included as a proxy measure of socioeconomic status, which can be associated with sleep. Health-related covariates included body mass index (BMI), intake of medication for vascular / heart problems, diabetes, smoking status, alcohol consumption status, and lifetime experience of depression. Intake of vascular medication was included as a proxy variable to take into account heart problems. Health-related variables were corrected for because they can independently impact on both sleep and brain structure. As additional sensitivity analyses, we re-ran all statistical tests 1) without lifetime depression covariate, and 2) without all health-related covariates. Please see supplementary sections S1.3 and S1.4 for definitions of each covariate variable, and for the details of the supplementary sensitivity analyses.

Of note, participants sleeping longer than recommended on average had higher BMI and were more likely to have self-reported diagnosis of diabetes, take medication for vascular or health problems, or have self-reported lifetime history of depression (supplementary sample information tables, all analyses with the main model). Participants sleeping less than recommended also had higher average BMI and were more likely to have self-reported diabetes or lifetime history of depression, but the differences in BMI and proportions of participants with health-related conditions were smaller. Participants sleeping both more or less than recommended came from areas with higher average Townsend deprivation indices.

Correction for false discovery rate (FDR)^36^ was performed by adjusting the *p*-values according to the procedure described in Benjamini, Heller & Yekutieli (2009),^37^ implemented within ‘mafdr’ function in MATLAB R2018a. Adjustment was performed separately for tests on each group of outcome variables and *p_FDR_* < 0.05 was taken as significant. Please see supplementary section S1.5 for the definition of outcome variable groups for correction for multiple comparisons.

## RESULTS

### Brain Structure Differences Associated with Sleeping Longer than Recommended

#### Overview of Brain Structure Differences

Sleep duration longer than the NSF recommendation was associated with differences in global brain measures, lobar and regional cortical thickness and volume measures. Differences were also found in fiber and tract-related MD, and the volume of the hippocampus. Please see top of Figure 1 for a Manhattan plot illustrating significant associations across all investigated brain measures.

**Figure 1.**
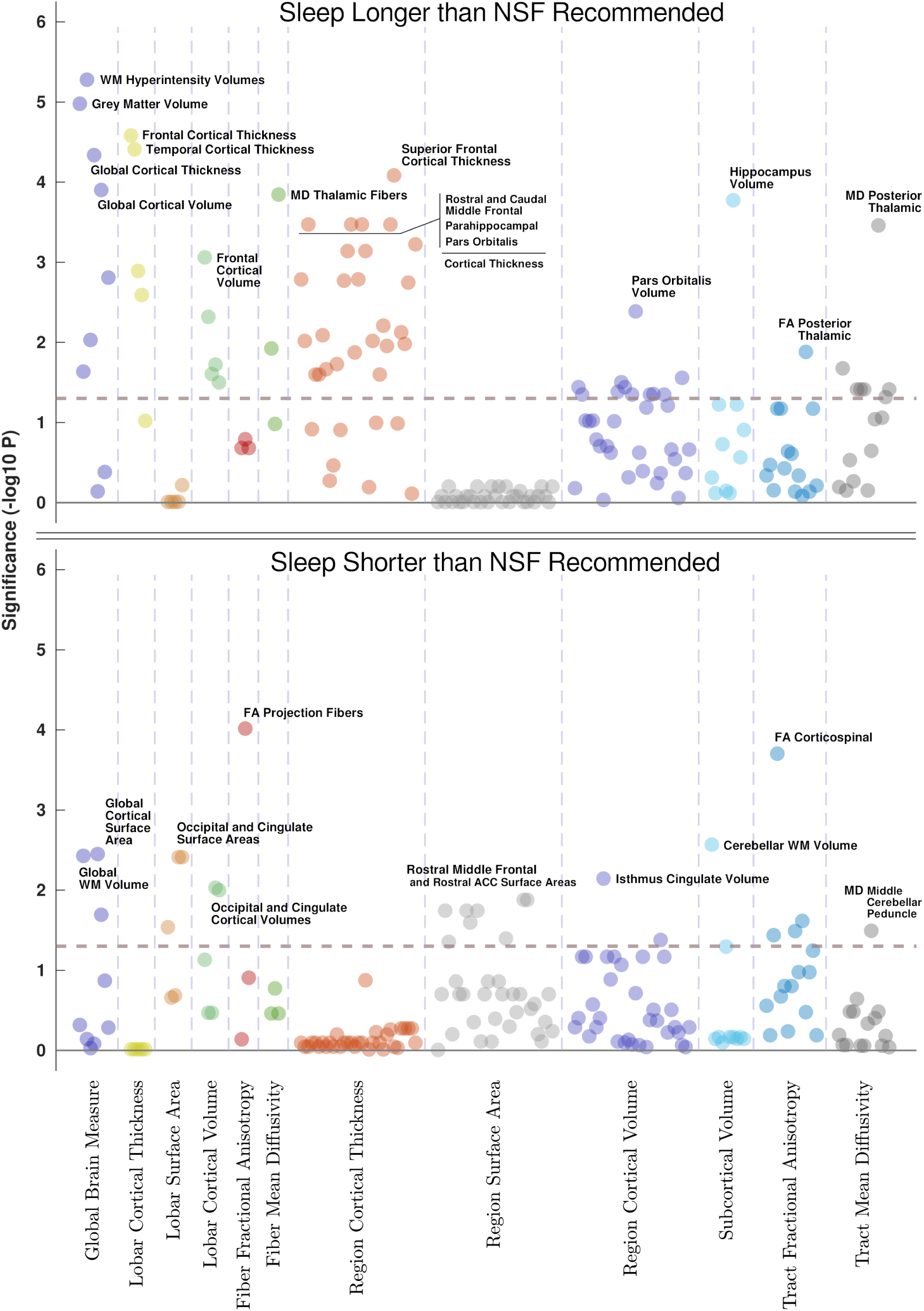
Manhattan plots illustrating significance values of associations of longer than recommended sleep duration (top), and shorter than recommended sleep duration (bottom) with brain structural measures. Dashed lines indicate statistical significance threshold (*p* = 0.05).

#### Global Brain Structure Differences

Sleeping longer than recommended was associated with lower overall grey and white matter volumes (*β_GM_* = −0.0207 *, p* < 0.0001 *; β_WM_* = −0.0136 *, p* = 0.0232), lower global cortical thickness and volume measures (*β_CT_* = −0.0229 *, p* < 0.0001 *; β_CV_* = −0.0129 *, p* = 0.000125), and higher brain age gap (*β_BAG_* = 0.0226 *, p* = 0.0093). Analyses also revealed poorer white matter structure, including higher volume of white matter hyperintensities (*β_WMHV_* = 0.0270 *, p* < 0.0001), and higher global MD and ISOVF measures (*β_gMD_* = 0.0188*, p* = 0.0016: *β_gISOVF_* = 0.0132*, p* = 0.0300). Left panel of Figure 2 illustrates effect sizes of significant associations of longer than recommended sleep with global brain measures.

**Figure 2.**
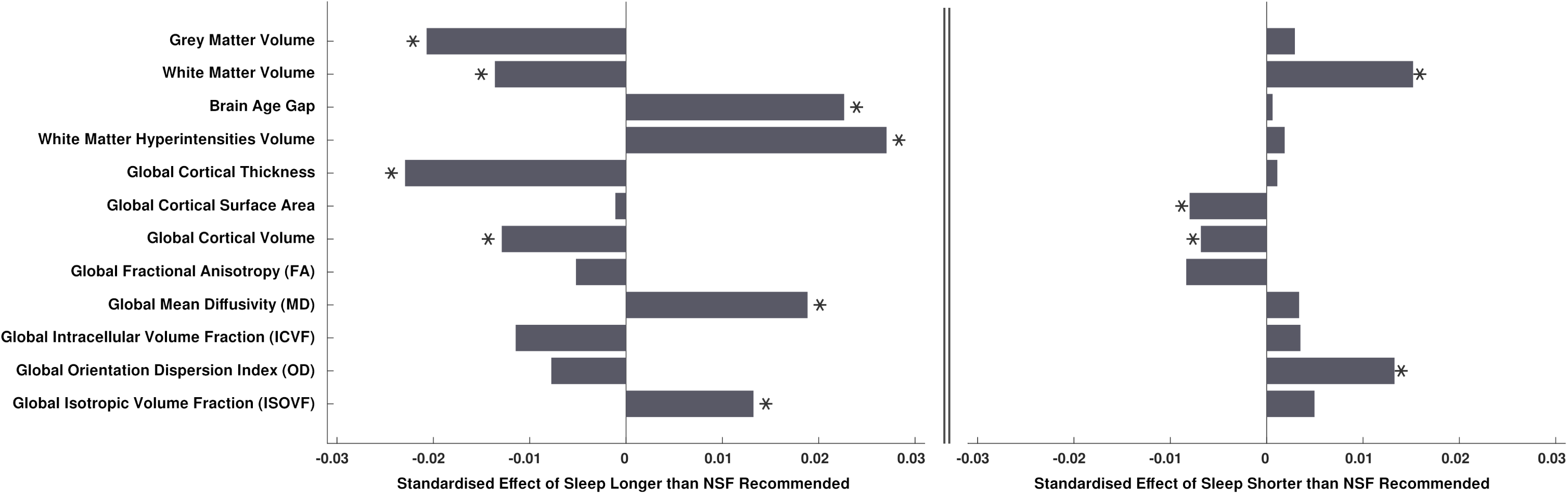
Barchart illustration of standardised effect sizes (beta coefficients) of associations of longer than recommended sleep duration (left), and shorter than recommended sleep duration (right) with global brain structural measures. Stars indicate statistically significant associations (*p*_FDR_ < 0.05).

#### Lobar and Regional Morphometry Differences

Lower cortical thickness was associated with sleeping longer than recommended in four out of five lobes (frontal *β_CT_* = −0.0255 *, p_FDR_* < 0.0001; temporal *β_CT_* = −0.0239 *, p_FDR_* < 0.0001; parietal *β_CT_* = −0.0191*, p_FDR_* = 0.0013; cingulate *β_CT_* = −0.0160 *, p_FDR_* = 0.0026), and in 25 out of 33 individual cortical regions. Largest associations were found in superior frontal cortex (*β_CT_* = −0.0256 *, p_FDR_* < 0.0001), rostral and caudal middle frontal areas (*β_CT_* = −0.0225 *, p_FDR_* = 0.0003 and *β_CT_* = −0.0222*, p_FDR_* = 0.0003), parahippocampal cortex ( *β_CT_* = −0.0216 *, p_FDR_* = 0.0003), pars orbitalis (*β_CT_* = −0.0215 *, p_FDR_* = 0.0003), and insula (*β_CT_* = −0.0208 *, p_FDR_* = 0.0006). Top left panels of Figures 3-4 and Figure 5 illustrate effect sizes related to lobar and regional cortical thickness measures.

**Figure 3.**
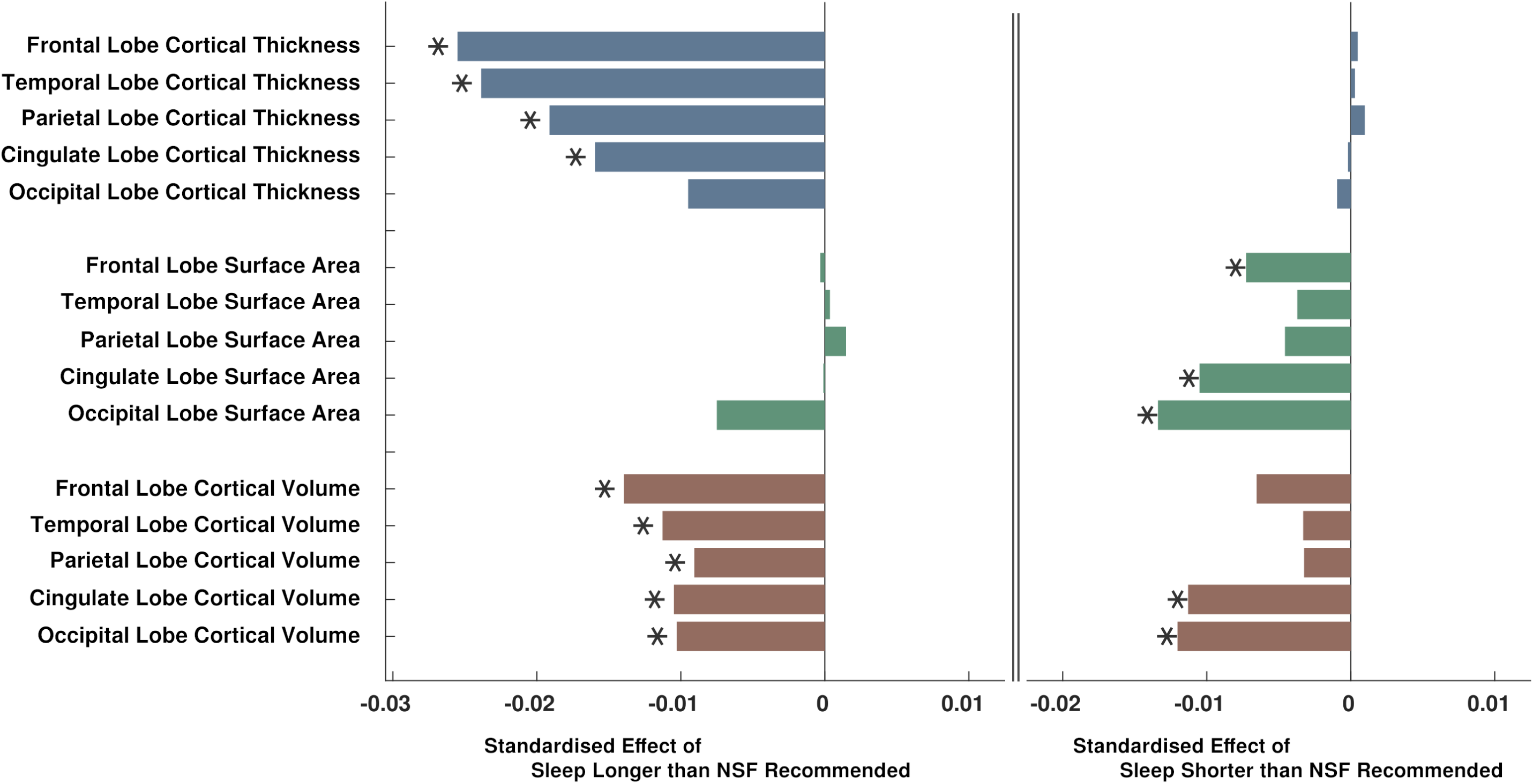
Barchart illustration of standardised effect sizes (beta coefficients) of associations of longer than recommended sleep duration (left), and shorter than recommended sleep duration (right) with lobar morphometric measures. Stars indicate statistically significant associations (*p*_FDR_ < 0.05).

**Figure 4.**
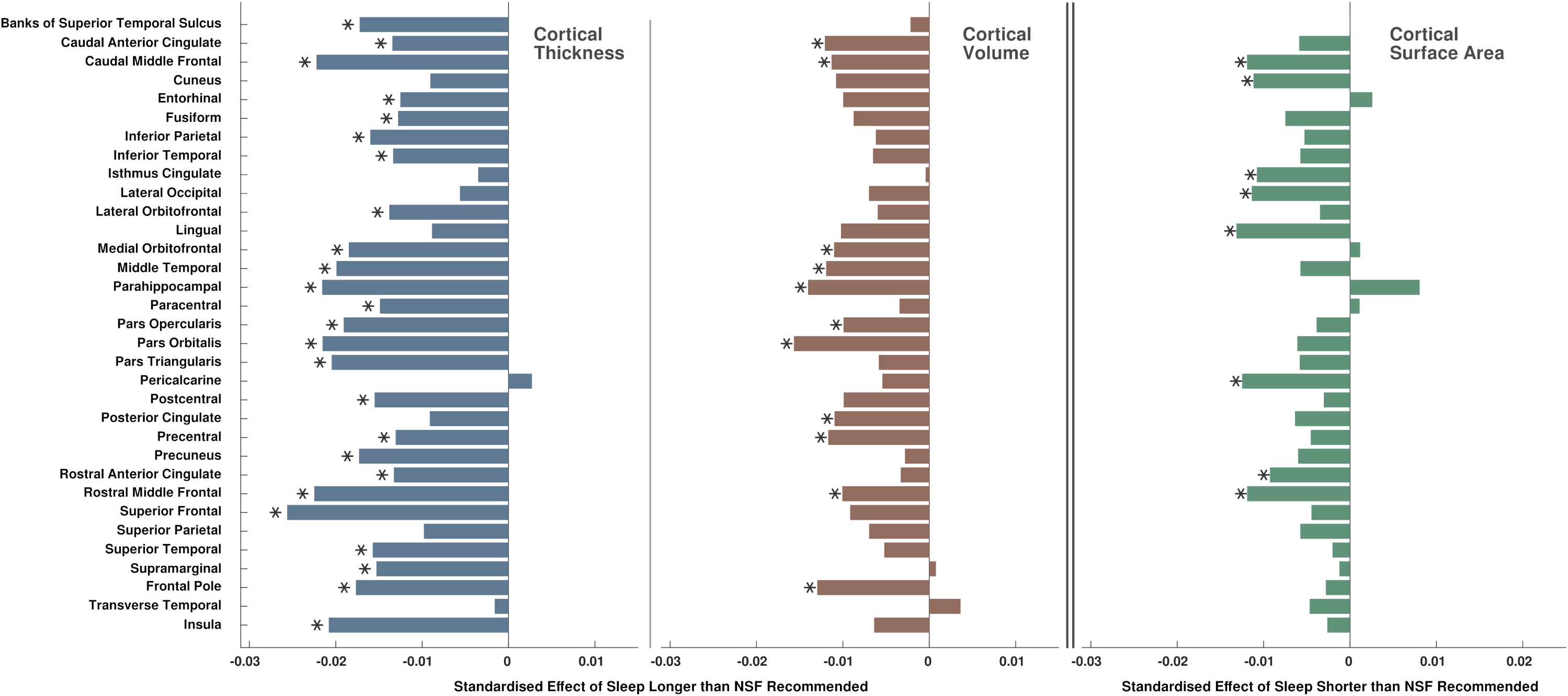
Barchart illustration of standardised effect sizes (beta coefficients) of associations of longer than recommended sleep duration with cortical thickness and cortical volume measures for individual regions (left and middle), and standardised effect sizes of associations of shorter than recommended sleep duration with cortical surface area measures (right). Stars indicate statistically significant associations (*p*_FDR_ < 0.05).

**Figure 5.**
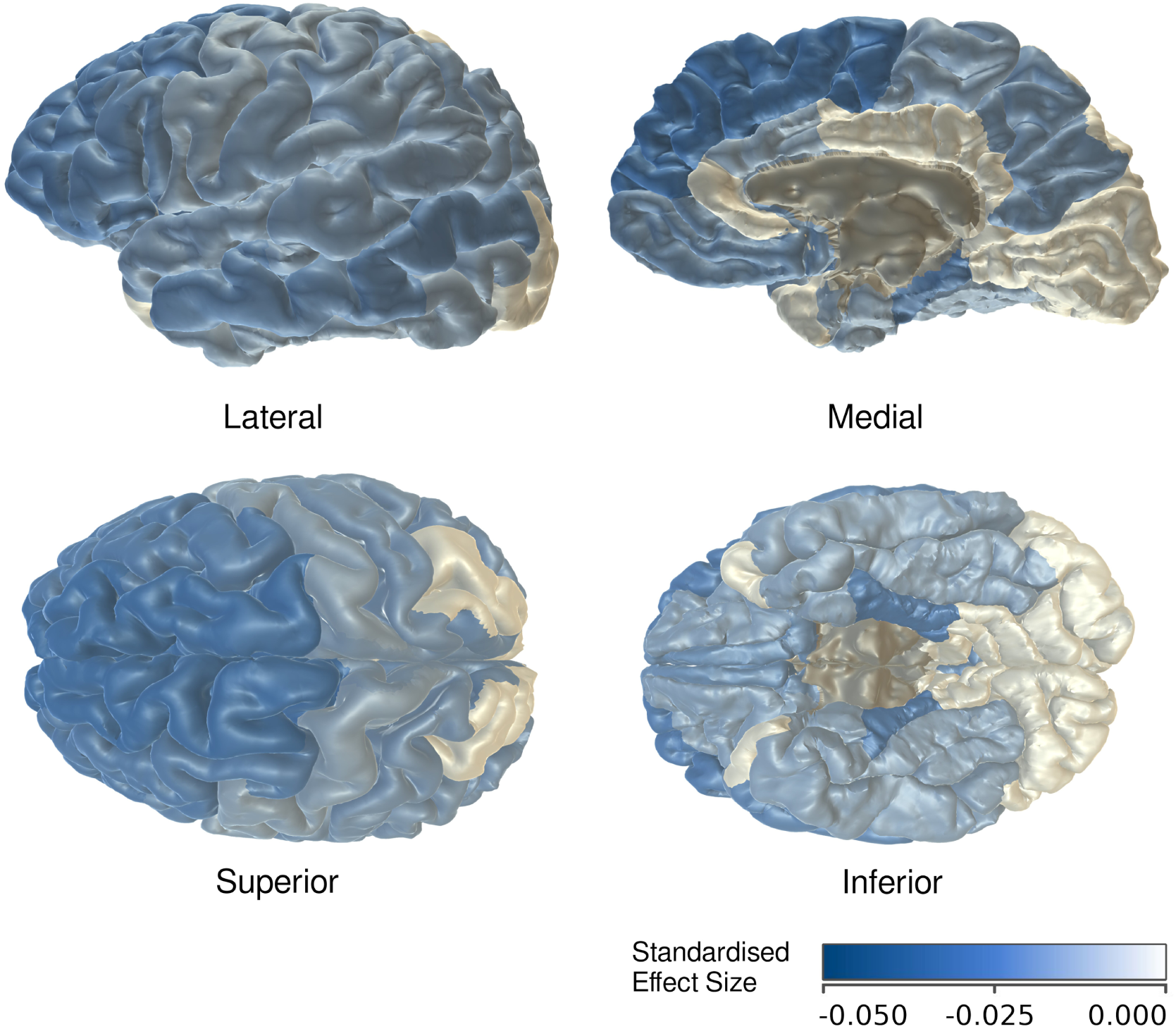
Heatmap illustration of lower cortical thickness in individual brain regions associated with sleeping longer than recommended.

Lower cortical volumes were found in all five lobes (frontal *β_CV_* = −0.0140 *, p_FDR_* = 0.0009; temporal *β_CV_* = −0.0113 *, p_FDR_* = 0.0048; parietal *β_CV_* = −0.0091 *, p_FDR_* = 0.0248; cingulate *β_CV_* = −0.0105 *, p_FDR_* = 0.0189; occipital *β_CV_* = −0.0103 *, p_FDR_* = 0.0316), and in 11 of 33 individual regions. Largest associations were found in pars orbitalis (*β_CV_* = −0.0157 *, p_FDR_* = 0.0041), parahippocampal (*β_CV_* = −0.0140 *, p_FDR_* = 0.0362), frontal pole (*β_CV_* = −0.0130 *, p_FDR_* = 0.0277), caudal anterior cingulate (*β_CV_* = −0.0120 *, p_FDR_* = 0.0362), and middle temporal regions (*β_CV_* = −0.0119 *, p_FDR_* = 0.0313). Figure 3 (bottom left) and Figure 4 (middle) illustrate effect sizes related to lobar and regional cortical volume measures.

#### Fiber and Tract White Matter Microstructure Differences

Sleeping longer than recommended was found to be associated with higher MD in association and thalamic fibers (*β_MD_* = 0.0162 *, p_FDR_* = 0.0119 and *β_MD_* = 0.0230 *, p_FDR_* = 0.0001), and in 7 of 15 individual tracts: posterior thalamic radiation (*β_MD_* = 0.0235 *, p_FDR_* = 0.0003), anterior thalamic radiation (*β_MD_* = 0.0164 *, p_FDR_* = 0.0211), superior thalamic radiation (*β_MD_* = 0.0125 *, p_FDR_* = 0.0482), forceps minor (*β_MD_* = 0.0164 *, p_FDR_* = 0.0385), inferior longitudinal fasciculus (*β_MD_* = 0.0144 *, p_FDR_* = 0.0385), inferior frontooccipital fasciculus (*β_MD_* = 0.0143 *, p_FDR_* = 0.0385), and uncinate fasciculus (*β_MD_* = 0.0135 *, p_FDR_* = 0.0385). Posterior thalamic radiation was found to also have lower FA (*β_FA_* = −0.0197 *, p_FDR_* = 0.0131).

#### Subcortical Volume Differences

Sleeping longer than recommended was found to be related to smaller hippocampal volume (*β* = −0.0201 *, p_FDR_* = 0.0002). No other associations with subcortical volumes were found.

### Brain Structure Differences Associated with Sleeping Less than Recommended

#### Overview of Brain Structure Differences

Sleeping less than the NSF recommended duration was associated with differences in global brain structure, lobar and regional cortical surface areas and volumes, fiber and tract-related FA, and the volume of the cerebellar white matter. Figure 1 (bottom) illustrates significant associations across all investigated brain measures.

#### Global Brain Structure Differences

Sleeping less than recommended was associated with higher overall white matter volume (*β_WM_* = 0.0152 *, p* = 0.0037), lower global cortical surface area (*β_SA_* = −0.0080 *, p* = 0.0035), lower global cortical volume (*β_CV_* = −0.0068 *, p* = 0.0202), and higher global OD (*β_gOD_* = 0.0133 *, p* = 0.0135).

Figure 2 (right) illustrates effect sizes of significant associations of shorter than recommended sleep with global brain measures.

#### Lobar and Regional Morphometry Differences

Sleeping less than recommended was associated with lower surface areas in the frontal lobe (*β_SA_* = −0.0073 *, p_FDR_* = 0.0290), cingulate gyrus (*β_SA_* = −0.0105 *, p_FDR_* = 0.0039), occipital lobe (*β_SA_* = −0.0134 *, p_FDR_* = 0.0039), and 8 of 33 individual cortical regions. Individual regions with smaller surface areas were the lingual gyrus (*β_SA_* = −0.0132 *, p_FDR_* = 0.0180), pericalcarine area (*β_SA_* = −0.0125 *, p_FDR_* = 0.0400), caudal and rostral middle frontal areas (*β_SA_* = −0.0119 *, p_FDR_* = 0.0180 and *β_SA_* = −0.0119 *, p_FDR_* = 0.0132), lateral occipital cortex (*β_SA_* = −0.0114 *, p_FDR_* = 0.0253), cuneus (*β_SA_* = −0.0112 *, p_FDR_* = 0.0442), isthmus cingulate (*β_SA_* = −0.0108 *, p_FDR_* = 0.0180), and rostral anterior cingulate (*β_SA_* = −0.0093 *, p_FDR_* = 0.0132). Figure 3 (right middle) and Figure 4 (right) illustrate effect sizes respectively related to lobar and regional surface areas.

Lower cortical volumes were found in the cingulate gyrus (*β_CV_* = −0.0113 *, p_FDR_* = 0.0093), occipital lobe (*β_CV_* = −0.0120 *, p_FDR_* = 0.01), and two individual regions: isthmus cingulate (*β_CV_* = −0.0146 *, p_FDR_* = 0.0072) and rostral anterior cingulate (*β_CV_* = −0.0094 *, p_FDR_* = 0.0418).

#### Fiber and Tract White Matter Microstructure Differences

Sleeping less than recommended was associated with lower FA in projection fibers (*β_FA_* = −0.0231 *, p_FDR_* = 0.0001), and in three individual white matter tracts – corticospinal (*β_FA_* = −0.0227 *, p_FDR_* = 0.0002), medial lemniscus (*β_FA_* = −0.0134 *, p_FDR_* = 0.0324), and parahippocampal cingulate (*β_FA_* = −0.0145 *, p_FDR_* = 0.0242). Conversely, higher FA was found in the cingulate gyrus (*β_FA_* = 0.0113*, p_FDR_* = 0.0364).

#### Subcortical Volume Differences

Sleeping less than recommended was related to higher cerebellar white matter volume (*β* = 0.0160 *, p_FDR_* = 0.0027). No other associations were identified.

### Brain Structure Differences Associated with Insomnia/Sleeplessness

Insomnia/sleeplessness was associated with higher global grey matter and white matter volumes (*β_GM_* = 0.0162 *, p* = 0.0049*; β_WM_* = 0.0198 *, p* = 0.0067), and with higher volumes of the putamen (*β* = 0.0222 *, p_FDR_* = 0.0016), hippocampus (*β* = 0.0206 *, p_FDR_* = 0.0016), and the amygdala (*β* = 0.0167 *, p_FDR_* = 0.0040).

## DISCUSSION

### Summary and Interpretation of Findings

#### Sleeping Longer than Recommended

We found that longer than NSF-recommended sleep duration was associated with lower overall grey matter and white matter volumes, and widespread lower cortical thickness and volume measures. Associations of cortical thickness were more pronounced compared to cortical volumes (Figure 3). Further, longer than recommended sleep was associated with advanced brain ageing, higher volume of white matter hyperintensities, higher diffusivity in the thalamic and association fibers (indicating potentially worse white matter integrity), and lower hippocampal volume.

Grey and white matter volumes and cortical thickness gradually decrease with age in healthy populations.^38^ Reductions in these measures have also been found in conditions which adversely affect both cognitive and emotional functions, including Alzheimer’s disease and dementia,^39–41^ cognitive impairment,^42^ depression^42–44^ and bipolar disorder.^45–49^ Increased diffusivity measures, indicating potential reductions in white matter integrity, have been observed after traumatic brain injury^50,51^ and in adult depression.^52^ White matter hyperintensities typically represent lesions that are a hallmark feature of cerebral small vessel disease.^53^ These lesions have also been found to be associated with depression,^42,54^ poorer cognition,^55^ and increased risk of cognitive impairment or stroke.^56–60^ Finally, the hippocampus has crucial roles in memory and learning.^61^ Decreased hippocampal volume has been reported with ageing,^62^ cognitive impairment,^63,64^ depression,^65,66^ and bipolar disorder.^48,67^ Taken together and in the context of the existing literature, our findings indicate that longer than recommended sleep duration is associated with advanced brain ageing and potentially worse brain health.

Of note, association of longer sleep with brain age gap was similar in magnitude to associations of overall grey matter volume, global cortical thickness and volume of white matter hyperintensities (Figure 2, left). This indicates that complex measures based on multiple aspects of brain structure may not be more sensitive to suboptimal sleep duration compared to simpler unimodal measures.

#### Sleeping Less than Recommended

Shorter than recommended sleep duration was related to higher global and cerebellar white matter volume, as well as lower global and regional cortical surface areas (primarily in the frontal, cingulate and occipital lobes) and lower FA (indicating potentially worse white matter integrity) in projection fibers.

Previous studies indicate that relatively higher global white matter volumes may be related to higher levels of physical activity and fitness.^68,69^ The cerebellum is crucially involved in movement control, as well as a number of other cognitive functions.^70^ It is clearly possible that shorter sleep times may lead to relatively higher levels of physical activity due to more time spent awake, which may in turn be linked to higher global and cerebellar white matter volumes.

Cortical surface area gradually decreases with age,^38^ with larger area measures associated with higher cognitive ability.^71^ Decreased surface area specifically of the isthmus cingulate (Figure 4, right) may reflect vulnerability to Alzheimer’s disease.^72^

Finally, lower FA of the projection fibers and the corticospinal tract (i.e. potentially reduced microstructural integrity) was previously identified in bipolar disorder, albeit in a small number of studies.^73^

Overall, our findings indicate that the pattern of differences in brain structure associated with sleeping less than recommended is quite different from that of sleeping more than recommended, and less indicative of poorer brain health.

#### Insomnia/Sleeplessness

Contrary to our initial hypothesis, we found that self-reported insomnia/sleeplessness was associated with higher (rather than lower) global grey and white matter volumes, with no evidence of changes in white matter microstructure. Further, we observed higher volumes of three subcortical structures: amygdala, hippocampus and putamen.

Global grey matter volume is correlated with general intelligence.^74^ The hippocampus is critical for cognitive function,^61^ and relatively higher hippocampal volumes are associated with higher levels of education^75^ and higher levels of physical activity.^76,77^ The putamen is a region likely to be involved in the circadian regulation of reward processing,^78^ and has previously been found to be enlarged in anxiety-related conditions and in bipolar disorder.^79,80^ Finally, the amygdala has critical roles in emotion and motivation,^81^ and higher amygdala volume has been identified in bipolar disorder,^82^ as well as genetic variants associated with risk for bipolar disorder.^83^

Overall, our findings indicate that self-reported insomnia/sleeplessness was related to higher volumes of brain structures putatively associated with intelligence, cognition, motivation and emotional processing. There is currently no robust evidence linking insomnia with higher intelligence, however our findings parallel previous results in the UK Biobank which indicated better cognitive performance in participants with past moderately-severe depression.^84^

### Study Strengths and Limitations

The two distinct advantages of our study are the large sample size and the wide range of structural brain measures, covering both grey and white matter. Previous studies of associations of sleep duration or insomnia and brain structure typically had *N* < 3000 and focused on relatively narrow sets of brain features. In contrast, the scope and power of the UK Biobank enabled sample sizes of between 9K and 33K (supplementary S1.6), and allowed us to uncover differences in brain structure distinctly associated with longer and shorter sleep, as well as insomnia/sleeplessness. Compared to previous work – including within the UK Biobank^85,86^ – we provide a more comprehensive characterisation of the brain structure associations with self-reported non-adherence to NSF guidelines and insomnia/sleeplessness.

Our study and results are complementary to those of Schiel et al. (2023),^87^ who have also investigated associations of regional grey matter volumes with longer or shorter sleep in the UK Biobank. To highlight the key differences, we studied regional volumes derived with the FreeSurfer toolkit rather than FAST, according to different brain atlases (Desikan-Killiany and Aseg vs. Harvard-Oxford).^88^ We also applied FDR rather than Bonferroni correction, grouped our statistical tests into smaller sets for correction (to facilitate independence between outcome variables, supplementary section S1.5), and combined data from left and right hemispheres as repeated measures to increase power. These differences most likely contributed to the higher numbers of significant associations found here as compared to Schiel et al. (2023). Together with Schiel et al. (2023),^87^ our study represents an example of how different and independent investigations can analyse non-overlapping subsets of extensive UK Biobank data with different methods in order to address similar research questions.

Despite these advantages, several limitations are acknowledged. Firstly, sleep variables were self-reported and may be subject to reporting biases. Future studies could confirm our findings by taking advantage of the actigraphy data collected within the UK Biobank from a subset of participants, albeit for the duration of only one week.^89^ Secondly, we did not assess causal directionality in the associations between sleep and brain structure measures. Future work could seek to apply causal inference methods such as Mendelian randomisation,^90^ potentially focussing on brain measures identified as significant here. Thirdly, we did not investigate changes in brain structure over time. Repeat brain imaging data collection is currently ongoing within the UK Biobank,^91^ and future data releases will enable high-powered longitudinal analyses. Fourthly, we did not correct for intake of medications that can have an effect on sleep, such as antidepressants, sedatives or hypnotics. Future studies can categorise the extensive self-reported medication data available within the UK Biobank and confirm if our findings hold when additionally correcting for psychotropic medication use. Finally, we did not address potential physiological mechanisms underlying the identified significant sleep and brain associations. These mechanisms could (at least in part) be addressed by using the rich proteomic data within the UK Biobank.^92^

### Conclusions

We made use of the breadth and scope of the UK Biobank dataset to identify that sleeping more than or less than the recommended NSF sleep time was associated with different patterns of altered brain structure. Specifically, longer than recommended sleep duration was related to differences in brain structural measures often associated with poorer brain health, while shorter sleep was associated with lower cortical surface areas. Future studies should seek to better characterise the mechanisms of these differences and define whether longer-than-recommended sleep duration represents a marker of clinical utility for population brain health.

## Supporting information

Sample Information

## Acknowledgements

The current study was funded and supported through the Lister Institute of Preventive Medicine award (reference 173096) and the Wellcome-University of Edinburgh Institutional Strategic Support Fund (reference 204804/Z/16/Z). LML is supported by a Royal College of Physicians of Edinburgh John, Margaret, Alfred and Stewart Sim Fellowship and a University of Glasgow Lord Kelvin / Adam Smith (LKAS) Fellowship. MJA and XS are supported by the Wellcome Trust (grant reference 220857/Z/20/Z). The research was conducted using the UK Biobank resource, with application number 4844. Provision of access to the UK Biobank was supported by the Wellcome Trust Strategic Award “Stratifying Resilience and Depression Longitudinally” (STRADL) (reference 104036/Z/14/Z). The current work has made use of the resources provided by the Edinburgh Compute and Data Facility (ECDF) (http://www.ecdf.ed.ac.uk/).

## Disclosure Statement

AMM previously received research grant support from Pfizer, Eli Lilly and Janssen, as well as speaker fees from Illumina. HCW previously received research grant support from Pfizer. None of these funding sources are connected to the present study. No other potential financial or non-financial conflicts of interest are reported.

## Data Availability Statement

The current study used data from the UK Biobank resource, which is available for health-related research upon registration and application through the UK Biobank Access Management System (https://www.ukbiobank.ac.uk/register-apply/).

## Notes

### Author Declarations

UK Biobank received ethical approval from the North West Multi-centre Research Ethics Committee in the UK (reference 11/NW/0382), and the current study received approval from the UK Biobank Access Committee (application #4844). All participants gave written informed consent.

### Summary of Updates

1) Added Table 1 with demographic and clinical characteristics of one representative studied sample (p. 31); 2) Added details of brain-predicted age computation methods within the main text (p. 6-7); 3) Moved paragraph outlining the differences between sleep-related participant groups in terms of clinical characteristics from supplementary document to the main text (p. 8); 4) Added reference to Benjamini, Heller & Yekutieli (2009) for the applied FDR correction procedure (p. 8); 5) Added discussion of the complementary UK Biobank study reported in Schiel et al. (2023) Brain Communications (p. 18); 6) Added mention of additional correction for psychotropic medication use as a direction for future work (p. 19); 7) Added details of the rationale for the selection of cortical and subcortical morphometric measures for the study (p. 5 in the supplementary document); 8) Added details of the rationale for grouping the outcome (brain measure) variables into sets for statistical correction (p. 10-11 in the supplementary document).

