## Supplementary Material for "Comprehensive Assessment of Sleep Duration, Insomnia and Brain Structure within the UK Biobank Cohort"

Aleks Stolicyn[1], Laura M. Lyall[2], Donald M. Lyall[2], Nikolaj Høier[1,3], Mark J. Adams[1], Xueyi Shen[1], James H. Cole[4,5], Andrew M. McIntosh[1], Heather C. Whalley[1], Daniel J. Smith[1]

1. Division of Psychiatry, Centre for Clinical Brain Sciences, University of Edinburgh, Edinburgh, United Kingdom

2. School of Health & Wellbeing, University of Glasgow, Glasgow, United Kingdom

3. Copenhagen Research Center for Mental Health CORE, Mental Health Center Copenhagen, Copenhagen University Hospital, Copenhagen, Denmark

4. Dementia Research Centre, Queen Square Institute of Neurology, University College London, United Kingdom

5. Centre for Medical Image Computing, Department of Computer Science, University College London, United Kingdom

Corresponding author:

Prof Daniel Smith
Division of Psychiatry, Centre for Clinical Brain Sciences, University of Edinburgh
Kennedy Tower, Royal Edinburgh Hospital, Morningside Park, Edinburgh EH10 5HF, UK.


### S1. MATERIALS AND METHODS

#### *S1.1 Sleep Measures*

The main analyses focused on three binary self-reported sleep-related exposure variables: 1) sleeping longer than recommended by the National Sleep Foundation (NSF) for adults[1]; 2) sleeping less than recommended by the NSF; and 3) insomnia/sleeplessness defined as trouble falling or staying asleep at night. The variables were based on the data collected as part of the UK Biobank touchscreen questionnaire and were defined as follows:

1) Sleep longer than recommended by the NSF – binary variable indicating whether participant's self-reported average sleep time per day is > 9 hours, as compared to 7-9 hours. Participants with average sleep times < 7 hours were excluded. Variable based on UKB data field 1160.

2) Sleep shorter than recommended by the NSF – binary variable indicating whether participant's self-reported average sleep time per day is < 7 hours, as compared to 7-9 hours. Participants with average sleep times > 9 hours were excluded. Variable based on UKB data field 1160.

3) Insomnia/sleeplessness – binary variable indicating self-reported usual trouble with falling asleep at night / waking up in the middle of the night, as compared to rarely or never having trouble with sleep. Participants who reported only occasional trouble with falling or staying asleep were excluded. Variable based on UKB data field 1200.

Supplementary analyses focused on four additional self-reported sleep-related variables: 1) sleep duration (in full hours per night); 2) contrast of longer than recommended sleep to shorter than recommended sleep; 3) chronotype preference (morning compared to evening); and 4) ease of getting up in the morning. The four additional variables were defined as follows:

1) Sleep duration – discrete variable indicating self-reported average number of hours slept during a typical 24 hour day, treated as a continuous variable in the analyses, based on UKB data field 1160.

2) Contrast of longer to shorter than recommended sleep – binary variable indicating whether participant's self-reported average sleep time per day is > 9 hours, as compared to < 7 hours. Participants with average sleep times 7-9 hours were excluded. Variable based on UKB data field 1160.

3) Late chronotype – binary variable indicating self-reported late chronotype (being an 'evening' person more so than a 'morning' person). Variable based on UKB data field 1180.

4) Ease of getting up – binary variable indicating self-reported 'fairly easy' or 'very easy' getting up in the morning, as compared to 'not very easy' or 'not at all easy'. Variable based on UKB data field 1170.

#### *S1.2 Brain Structure Measures*

##### S1.2.1 Global Brain Measures

Global grey and white matter volumes (normalised for head size), as well as hyperintensity volumes were derived by the UK Biobank respectively with the SIENAX and BIANCA toolkits (UKB data fields 25005, 25007 and 25781).[2,3]

Global cortical thickness and surface area measures for the two hemispheres were derived by the UK Biobank with FreeSurfer version 6.0[4] (UKB data fields 26721, 26755, 26822 and 26856). Global cortical volume measures were derived for each hemisphere locally as the sums of 33 regional hemispheric cortical volumes (UKB data fields 26789-26821 and 26890-

26922).

Brain age gap measure was derived locally for a subset of UK Biobank participants ($N$ = 20895) as the differences between age estimated from T1 scan (brain-predicted age) and actual chronological age, additionally residualised for chronological age. Brain-predicted age measures were derived with brainageR toolkit[5] (version 2.1 available at https://github.com/james-cole/brainageR). brainageR toolkit applies Gaussian processes regression to estimate age from normalised voxel-wise measures of grey matter, white matter and cerebrospinal fluid obtained from T1-weighted brain scans.

Global white matter microstructure measures – fractional anisotropy (FA), mean diffusivity (MD), intra-cellular volume fraction (ICVF), isotropic volume fraction (ISOVF), and orientation dispersion index (OD) – were derived as the scores on the first unrotated principal components from principal component analyses (PCA) which combined relevant measures (FA, MD, ICVF, ISOVF or OD) from 15 individual white matter tracts (please see section S1.2.3 below). Briefly, FA measures coherence of water molecule movement along white matter tracts, MD measures free water molecule diffusion in all directions, ICVF indexes white matter neurite density, ISOVF measures the fractions of white matter tracts with freely moving water, and OD measures overall disorganisation of the tracts.[3,6,7] Higher FA and ICVF indicate potentially better white matter microstructural organisation, while higher MD, ISOVF and OD indicate potentially worse white matter integrity,[6,7] although please see Jones et al. (2013) for further discussion of these measures.[8]

##### S1.2.2 Lobar and Fiber Measures

Lobar cortical thickness, surface area and volume measures were derived for five bilateral lobes – frontal, temporal, cingulate, parietal and occipital. Lobar cortical thickness measures were calculated as sums of relevant (lobe-assigned) regional thickness measures weighted by respective ratios of regional surface area to overall lobar surface area. Lobar surface area and volume measures were calculated as sums of relevant regional surface area or volume measures. Please see Table S1 for a mapping of individual cortical regions to the five lobes.

Fiber-related FA, MD, ICVF, OD and ISOVF were derived for three fiber bundles (association, projection and thalamic) as the scores on the first unrotated principal components from PCA analyses which combined measures of individual white matter tracts belonging to the bundle. Please see Table S2 for a mapping of individual white matter tracts onto fiber bundles. Please see Tables S3-S7 for the sets of loadings of individual white matter tract measures onto fiber bundle measures.

###### S1.2.3 Regional Brain Measures

Regional grey matter morphometric data consisted of cortical thickness, surface area and volume measures for 33 individual bilateral regions defined according to the Desikan-Killiany atlas, as well as volumes of 10 bilateral subcortical regions defined according to the FreeSurfer Aseg atlas (UKB data fields 26722-26922 in category 192, and data fields 26556-26565 and 26587-26596 in category 190), derived by the UK Biobank with FreeSurfer version 6.0.[3,4] All cortical morphometric measures in category 192 were studied because they cover the entire cerebral cortex and do not overlap. The 10 subcortical regions (the amygdala, the hippocampus, nucleus accumbens, caudate, putamen, pallidum, thalamus, ventral diencephalon, and the cerebellar grey and white matter) were selected on the basis that these regions have important roles in cognition, memory, emotion and movement,[9–11] and are of most interest to study in relation to sleep disruption.[12] These subcortical regions were investigated in relation to cognition and mood disorder in the past studies in our lab.[13–15]

White matter microstructure measures (FA, MD, ICVF, OD and ISOVF) for 12 bilateral and 3 unilateral individual tracts, defined according to AutoPtx atlas,[3,16] were derived with the probabilistic tractography toolkit in FSL (BEDPOSTx / PROBTRACKx) by the UK Biobank (UKB data fields 25488-25541, 25650-25703, 25704-25730).[3,17,18]

**S1.2.4 Outlier Definitions**

Brain structural measure values were considered outliers if they were more than the set number of standard deviations (SD) different from the sample mean for that measure. Brain measures were grouped into 9 sets and participants were excluded from analyses for any set where they had an outlier value for at least one measure in that set. The brain measure sets and outlier thresholds were following:

(1) **Global grey and white matter volumes** (two measures) – 3.5 SD outlier threshold;

(2) **Overall volume of white matter hyperintensities** (single measure) – 3.5 SD outlier threshold;

(3) **Brain age gap** (single measure) – 3.5 SD outlier threshold;

(4) **Global and lobar morphometry** (36 measures in total – 6 bilateral cortical thickness / 6 bilateral surface area / 6 bilateral cortical volume) – 3.5 SD outlier threshold;

(5) **Cortical and subcortical regional morphometry** (218 measures in total – 33 bilateral cortical thickness / 33 bilateral surface area / 33 bilateral cortical volume / 10 bilateral subcortical volume) – 5 SD outlier threshold;

(6) **Global and fiber-related DTI white matter microstructure** (8 measures in total – 4 FA / 4 MD) – 4.5 SD outlier threshold;

(7) **Tract-related DTI white matter microstructure** (54 measures in total – 3 unilateral FA / 12 bilateral FA / 4 unilateral MD / 12 bilateral MD) – 5 SD outlier threshold;

(8) **Global and fiber-related NODDI (neurite orientation and dispersion density imaging) white matter microstructure** (12 measures in total – 4 ICVF / 4 OD / 4 ISOVF) – 4.5

SD outlier threshold;

(9) **Tract-related NODDI white matter microstructure** (81 measures in total – 3 unilateral ICVF / 12 bilateral ICVF / 3 unilateral OD / 12 bilateral OD / 3 unilateral ISOVF / 12 bilateral ISOVF) – 5 SD outlier threshold.

Outlier criteria were specified to enable optimal numbers of excluded participants for each of the above sets of brain measures.

##### *S1.3 Covariate Variables*

Analyses of all unilateral brain measures were performed with generalized linear regression models (GLM) with the following model structure:

Brain measure (Outcome) ~ Sleep measure (Exposure) + Age + Sex + Ethnic background +
Townsend index + Body mass index (BMI) +
Vascular medication status + Diabetes diagnosis +
Smoking status + Alcohol consumption status +
Lifetime depression status + Within-scanner head coordinates

Analyses of cortical thickness, surface area and volume measures included an additional covariate term for intracranial volume (ICV), standardised across the entire available sample.

Analyses of all bilateral brain measures were performed with linear mixed effects (LME) models with the same structure as above and an additional term (Hemisphere | Participant) to treat data from left and right hemispheres as repeated measures.

Covariate variables were defined as follows:

**Age / Sex / Townsend index / BMI** – obtained respectively from UKB data fields 21003, 31, 189 and 21001.

**Ethnic background –** binary variable indicating 'White British' as compared to any other ethnicity (due to predominance of white British participants in the UK Biobank),[19] based on UKB data field 21000.

---

**Vascular medication status –** binary variable indicating intake of medication relevant for vascular or heart problems, used as a proxy to control for heart problems, based on UKB data fields 6177 and 6153.

---

**Diabetes diagnosis –** binary variable indicating self-reported diagnosis of diabetes from doctor's advice, based on UKB data field 2443.

---

**Smoking status –** binary variable indicating current or previous smoking status (as compared to never having smoked), based on UKB data field 20116.

---

**Alcohol consumption status –** binary variable indicating alcohol consumption habits (once a week or more often as compared to less than once a week), based on UKB data field 1558.

---

**Lifetime depression status –** binary variable capturing lifetime experience of depression, based on UKB data fields 4598, 4609, 2090, 4631, 5375 and 2100. Here lifetime experience of depression was defined as follows: 1) Participant reported ever seeing a doctor (GP) or a psychiatrist for depression, anxiety or nerves (UKB data fields 2090 and 2100), *and* either 2A) Participant reported ever feeling depressed for a whole week (UKB data field

4598) with the longest experienced period of depression of at least two weeks (UKB data field 4609), *or* 2B) Participant reported ever experiencing anhedonia (lack of interest in daily activities) for a whole week (UKB data field 4631) with the longest experienced period of anhedonia (disinterest) of at least two weeks (UKB data field 5375).

---

**Within-scanner head coordinates –** three coordinates defining participant head position within the scanner (included to account for potential differences in magnetic field), based on UKB data fields 25756-25758.

---

**ICV –** total intractranial volume estimated with FreeSurfer, obtained from UKB data field 26521.

---

##### *S1.4 Sensitivity Analyses*

Sensitivity analyses included rerunning all statistical tests for the three main exposure variables 1) without the lifetime depression covariate (reduced model I), and 2) without all health-related covariates (reduced model II). Tested statistical models without the lifetime depression covariate had the following structure:

Brain measure (Outcome) ~ Sleep measure (Exposure) + Age + Sex + Ethnic background + Townsend index + Body mass index (BMI) + Vascular medication status + Diabetes diagnosis + Smoking status + Alcohol consumption status + Lifetime depression status + Within-scanner head coordinates

Tested statistical models without all health-related covariates had the following structure:

Brain measure (Outcome) ~ Sleep measure (Exposure) + Age + Sex + Ethnic background + Townsend index + Within-scanner head coordinates

##### *S1.5 Multiple Comparisons Correction*

Correction for false discovery rate (FDR) was performed separately for tests within 17 groups of outcome variables, separately for each investigated sleep measure (exposure). The outcome variable groups for FDR correction were following:

1) 5 lobar cortical thickness measures;
2) 5 lobar surface area measures;
3) 5 lobar cortical volume measures;
4) 33 regional cortical thickness measures;
5) 33 regional surface area measures;
6) 33 regional cortical volume measures;
7) 10 subcortical volume measures;
8) 3 fiber-related FA measures;
9) 3 fiber-related MD measures;
10) 3 fiber-related ICVF measures;
11) 3 fiber-related OD measures;
12) 3 fiber-related ISOVF measures;
13) 15 individual tract-related FA measures;
14) 15 individual tract-related MD measures;
15) 15 individual tract-related ICVF measures;
16) 15 individual tract-related OD measures;
17) 15 individual tract-related ISOVF measures.

The above outcome variable groups were defined so that brain measures in each group were of the same type but related to different brain areas, in order to maximise independence between the tests within each group. Some of the measures across groups cannot be considered independent – for example, cortical volume of a particular region depends both on that region's surface area and thickness, white-matter measures (FA / MD / ICVF / OD / ISOVF) capture different but inter-related aspects of microstructure, while lobar and fiber-related measures depend directly on the constituent regional and tract-related measures. By performing FDR correction separately within groups of outcome variables defined by type, we aimed to achieve a better balance between the possibilities of type I and type II errors, as compared to correcting across all of the statistical tests taken together. A similar approach (albeit with fewer measure types) was taken in previous studies in our lab.[20–22]

FDR correction was not performed on the tests of global brain measures because these measures were related to different but inter-related aspects of global brain structure, and were not considered to belong to a single family.

##### *S1.6 Sample Characteristics*

Participants were excluded in a way so as to maximise sample sizes in all analyses. Participants were excluded from any analysis where they had missing sleep-related (exposure) or covariate data. Participants were also excluded from analyses for any particular set of outcome (brain) variables if they had a missing value or an outlier value for any variable in that set (please see section S1.2.4 above for the definition of outcome variable sets and outlier values). There were therefore 162 UK Biobank subsamples analysed in total (9 brain measure sets × 3 covariate sets × 6 sleep measures), with sample sizes ranging between 9K and 36K participants (up to 32K in the analyses with the main model / covariate set). Please see the accompanying supplementary CSV tables for exact sample sizes and summary demographic characteristics for each analysed participant sample. For analyses where the exposure (sleep-related) variable was binary, summary sample characteristics are presented separately for cases and controls, where participants with positive exposure values were considered cases.

Demographic and health-related characteristics were compared between cases and controls with two-sample *t*-tests (for continuous variables such as age or BMI) or with chi-squared tests (for participant proportions e.g. related to health conditions). CSV tables with summary demographic characteristics are grouped according to the analysed exposure variable, with the filename of each table denoting 1) the analysed exposure variable (section S1.1), 2) the outcome variables set (section S1.2.4), and 3) the set of covariates (main or reduced model, section S1.4).

### S2. RESULTS

#### *S2.1 Associations of Main Investigated Sleep Measures with Fiber and Tract-related NODDI Measures*

##### S2.1.1 Fiber and Tract-related NODDI Differences Associated with Sleep Longer than Recommended

Sleeping longer than recommended was associated with lower ICVF and OD in thalamic fibers ($\beta = -0.0149,\ p_{FDR} = 0.0499$ and $\beta = -0.0207,\ p_{FDR} = 0.0017$), lower OD in projection fibers ($\beta = -0.0166,\ p_{FDR} = 0.0130$), and higher ISOVF in thalamic fibers ($\beta = 0.0180,\ p_{FDR} = 0.0063$).

With regard to individual white matter tracts, lower OD was found in superior thalamic radiation ($\beta = -0.0240,\ p_{FDR} = 0.0006$) and corticospinal tract ($\beta = -0.0195,\ p_{FDR} = 0.0071$), while higher ISOVF was found in posterior thalamic radiation ($\beta = 0.0165,\ p_{FDR} = 0.0440$).

##### S2.1.2 Fiber and Tract-related NODDI Differences Associated with Sleep Shorter than Recommended

Sleeping less than recommended was associated with higher OD in projection fibers

($\beta = 0.0210$, $p_{FDR} = 0.0005$), and in three individual tracts – medial lemniscus ($\beta = 0.0242$, $p_{FDR} < 0.0001$), corticospinal tract ($\beta = 0.0213$, $p_{FDR} = 0.0003$), and superior longitudinal fasciculus ($\beta = 0.0124$, $p_{FDR} = 0.0346$). Lower ISOVF was found in middle cerebellar peduncle ($\beta = -0.0185$, $p_{FDR} = 0.0151$), but higher ISOVF was found in superior thalamic radiation ($\beta = 0.0141$, $p_{FDR} = 0.0185$).

###### S2.1.3 Fiber and Tract-related NODDI Differences Associated with Insomnia/Sleeplessness

Insomnia/sleeplessness was associated with higher OD in projection fibers ($\beta = 0.0186$, $p_{FDR} = 0.0489$). No other fiber or tract-related NODDI measure differences were identified.

##### *S2.2 Brain Structure Associations of Additional Sleep Measures*

###### S2.2.1 Brain Structure Differences Associated with *Sleep Duration*

At the global level, longer sleep duration was associated with lower overall grey matter and white matter volumes (respectively $\beta = -0.0110$, $p = 0.0079$ and $\beta = -0.0131$, $p = 0.0123$), lower overall cortical thickness ($\beta = -0.0187$, $p = 0.0001$), lower global OD ($\beta = -0.0136$, $p = 0.011$), and higher brain age gap ($\beta = 0.0176$, $p = 0.0203$). Further to that, longer sleep appeared to be associated with higher overall cortical surface area ($\beta = 0.0068$, $p = 0.0126$). Associations related to grey and white matter volumes, cortical thickness and brain age gap appeared consistent in their direction and magnitude with differences associated with sleeping longer than recommended (please see the results section of the main text).

At the lobar level, longer sleep duration was associated with lower cortical thickness in frontal ($\beta = -0.0196$, $p_{FDR} = 0.0002$), temporal ($\beta = -0.0198$, $p_{FDR} = 0.0002$), parietal ($\beta = -0.0139$, $p_{FDR} = 0.0066$), and cingulate ($\beta = -0.0131$, $p_{FDR} = 0.0063$) lobes, and with higher cingulate surface area ($\beta = 0.0110$, $p_{FDR} = 0.0034$).

At the regional level, longer sleep duration was associated with lower cortical thickness in 22 out of 33 individual regions, with largest effect sizes seen in pars triangularis ($\beta = -0.0279$, $p_{FDR} < 0.0001$), rostral middle frontal ($\beta = -0.0243$, $p_{FDR} < 0.0001$), superior temporal ($\beta = -0.0242$, $p_{FDR} < 0.0001$), superior frontal ($\beta = -0.0217$, $p_{FDR} < 0.0001$) and frontal polar ($\beta = -0.0208$, $p_{FDR} < 0.0001$) regions. Higher surface areas were observed in rostral anterior cingulate ($\beta = 0.0110$, $p_{FDR} = 0.0019$), isthmus cingulate ($\beta = 0.0108$, $p_{FDR} = 0.0362$), and transverse temporal ($\beta = 0.0078$, $p_{FDR} = 0.0476$) regions.

With regard to subcortical structures, longer sleep duration was associated with lower volumes of the cerebellar white matter ($\beta = -0.0151$, $p_{FDR} = 0.0051$), hippocampus ($\beta = -0.0134$, $p_{FDR} = 0.0051$), putamen ($\beta = -0.0121$, $p_{FDR} = 0.0142$) and thalamus ($\beta = -0.0111$, $p_{FDR} = 0.0069$).

With regard to fiber-related white matter microstructure, longer sleep duration was associated with higher FA and MD measures in projection fibers (respectively $\beta = 0.0264$, $p_{FDR} < 0.0001$ and $\beta = 0.0189$, $p_{FDR} = 0.0014$), with lower OD measures in projection and thalamic fibers (respectively $\beta = -0.0247$, $p_{FDR} < 0.0001$ and $\beta = -0.0150$, $p_{FDR} = 0.0064$), and with higher ISOVF in projection fibers ($\beta = 0.0197$, $p_{FDR} = 0.0012$).

With regard to tract-related white matter microstructure, longer sleep duration was associated with lower FA in the cingulate gyrus ($\beta = -0.0136$, $p_{FDR} = 0.0109$), higher FA in corticospinal tract ($\beta = 0.0277$, $p_{FDR} < 0.0001$), superior thalamic radiation ($\beta = 0.0169$, $p_{FDR} = 0.0109$) and medial lemniscus ($\beta = 0.0123$, $p_{FDR} = 0.0452$), and higher MD in middle cerebellar peduncle ($\beta = 0.0248$, $p_{FDR} = 0.0001$), forceps minor ($\beta = 0.0200$, $p_{FDR} = 0.0017$) and corticospinal tract ($\beta = 0.0139$, $p_{FDR} = 0.0410$). Lower ICVF was found in superior thalamic radiation ($\beta = -0.0159$, $p_{FDR} = 0.0241$), forceps minor ($\beta = -0.0151$, $p_{FDR} = 0.0302$), cingulate gyrus ($\beta = -0.0149$, $p_{FDR} = 0.0241$), and uncinate fasciculus ($\beta = -0.0138$, $p_{FDR} = 0.0302$). Lower OD was found in corticospinal tract ($\beta = -0.0316$, $p_{FDR} < 0.0001$), medial lemniscus ($\beta = -0.0230$, $p_{FDR} < 0.0001$), superior thalamic radiation ($\beta = -0.0191$, $p_{FDR} < 0.0001$), superior longitudinal fasciculus ($\beta = -0.0146$, $p_{FDR} = 0.0053$), and inferior longitudinal fasciculus ($\beta = -0.0119$, $p_{FDR} = 0.0391$). Higher OD was found in cingulate gyrus ($\beta = 0.0121$, $p_{FDR} = 0.0173$). Lower ISOVF was found in cingulate gyrus ($\beta = -0.0178$, $p_{FDR} < 0.0001$) and superior thalamic radiation ($\beta = -0.0175$, $p_{FDR} < 0.0001$). Higher ISOVF was found in middle cerebellar peduncle ($\beta = 0.0254$, $p_{FDR} < 0.0001$).

###### S2.2.2 Brain Structure Differences Associated with *Longer than Recommended Sleep Compared to Shorter than Recommended Sleep*

Overall, associations of longer-than-recommended compared to shorter-than-recommended sleep (longer-to-shorter sleep contrast) were similar to those of longer than recommended sleep (please see the results section of the main text).

At the global level, longer-to-shorter sleep contrast was associated with lower overall grey and white matter volumes (respectively $\beta = -0.0348,\ p < 0.0001$ and $\beta = -0.0281,\ p = 0.0067$), lower global cortical thickness and volume measures (respectively $\beta = -0.0377,\ p = 0.0001$ and $\beta = -0.0182,\ p = 0.0018$), and higher brain age gap ($\beta = 0.0404,\ p = 0.0078$). With regard to global white matter, longer-to-shorter sleep contrast was associated with higher volume of white matter hyperintensities ($\beta = 0.0456,\ p < 0.0001$), higher global MD ($\beta = 0.0334,\ p = 0.0012$), and lower global ICVF ($\beta = -0.0245,\ p = 0.0271$).

At the lobar level, longer-to-shorter sleep contrast was associated with lower cortical thickness in frontal lobe ($\beta = -0.0417,\ p_{FDR} = 0.0001$), temporal lobe ($\beta = -0.0397,\ p_{FDR} = 0.0001$), parietal lobe ($\beta = -0.0301,\ p_{FDR} = 0.0044$), and cingulate cortex ($\beta = -0.026,\ p_{FDR} = 0.0051$). Lower volumes were found in frontal and temporal lobes (respectively $\beta = -0.0206,\ p_{FDR} = 0.0065$ and $\beta = -0.0164,\ p_{FDR} = 0.0252$).

At the cortical regional level, longer-to-shorter sleep contrast was associated with lower cortical thickness in 25 out of 33 individual regions, with largest effect sizes seen in superior frontal cortex ($\beta = -0.042,\ p_{FDR} = 0.0002$), pars triangularis ($\beta = -0.0415,\ p_{FDR} = 0.0002$), rostral and caudal middle frontal areas (respectively $\beta = -0.0413,\ p_{FDR} = 0.0002$ and $\beta = -0.0364,\ p_{FDR} = 0.0008$), and pars orbitalis ($\beta = -0.038,\ p_{FDR} = 0.0004$). Lower volumes were found for pars orbitalis ($\beta = -0.024,\ p_{FDR} = 0.0256$) and frontal pole ($\beta = -0.0213,\ p_{FDR} = 0.049$).

Similarly to sleeping longer than recommended, longer-to-shorter sleep contrast was found to be related to smaller hippocampal volume ($\beta = -0.0359, \ p_{FDR} < 0.0001$), but no other associations with subcortical volumes were found.

With regard to fiber-related white matter microstructure, longer-to-shorter sleep contrast was associated with higher FA in projection fibers ($\beta = 0.0277, \ p_{FDR} = 0.0352$), higher MD in association ($\beta = 0.0279, \ p_{FDR} = 0.013$), projection ($\beta = 0.0237, \ p_{FDR} = 0.0273$) and thalamic ($\beta = 0.0397, \ p_{FDR} = 0.0002$) fibers, also lower ICVF in association ($\beta = -0.0244, \ p_{FDR} = 0.0427$) and thalamic ($\beta = -0.0294, \ p_{FDR} = 0.0209$) fibers, lower OD in projection ($\beta = -0.0368, \ p_{FDR} = 0.0012$) and thalamic ($\beta = -0.0408, \ p_{FDR} = 0.0003$) fibers, and higher ISOVF in thalamic fibers ($\beta = 0.0274, \ p_{FDR} = 0.0203$).

With regard to tract-related white matter microstructure, longer-to-shorter sleep contrast was associated with higher FA in corticospinal tract and superior thalamic radiation (respectively $\beta = 0.0326, \ p_{FDR} = 0.014$ and $\beta = 0.0295, \ p_{FDR} = 0.0374$), and lower FA in posterior thalamic radiation ($\beta = -0.032, \ p_{FDR} = 0.014$). Higher MD was found in 8 of 15 individual tracts: anterior and posterior thalamic radiations (respectively $\beta = 0.0321, \ p_{FDR} = 0.0045$ and $\beta = 0.0384, \ p_{FDR} = 0.0012$), forceps minor ($\beta = 0.0361, \ p_{FDR} = 0.0045$), inferior frontooccipital fasciculus ($\beta = 0.0263, \ p_{FDR} = 0.0245$), inferior longitudinal fasciculus ($\beta = 0.0259, \ p_{FDR} = 0.0245$), middle cerebellar peduncle ($\beta = 0.0253, \ p_{FDR} = 0.0409$), uncinate fasciculus ($\beta = 0.0249, \ p_{FDR} = 0.0245$), superior thalamic radiation ($\beta = 0.0226, \ p_{FDR} = 0.0385$).

With regard to tract-related NODDI measures, lower ICVF was found in forceps major ($\beta = -0.0309$, $p_{FDR} = 0.0473$) and in posterior thalamic radiation ($\beta = -0.0309$, $p_{FDR} = 0.0473$). Lower OD was found in superior thalamic radiation ($\beta = -0.0461$, $p_{FDR} = 0.0001$) and corticospinal tract ($\beta = -0.0417$, $p_{FDR} = 0.0004$).

##### S2.2.3 Brain Structure Differences Associated with *Late Chronotype*

At the global level, late chronotype was associated with lower overall white matter volume ($\beta = -0.0130$, $p = 0.0130$), lower overall surface area ($\beta = -0.0104$, $p = 0.0001$) and lower global ICVF ($\beta = -0.0126$, $p = 0.0235$).

At the lobar lever, late chronotype was associated with lower surface areas of frontal ($\beta = -0.0068$, $p_{FDR} = 0.0393$), temporal ($\beta = -0.0080$, $p_{FDR} = 0.0198$), parietal ($\beta = -0.0156$, $p_{FDR} < 0.0001$) and cingulate ($\beta = -0.0070$, $p_{FDR} = 0.0393$) lobes, and with lower parietal lobe volume ($\beta = -0.0089$, $p_{FDR} = 0.0423$).

At the regional level, late chronotype was associated with higher cortical thickness of the cuneus ($\beta = 0.0186$, $p_{FDR} = 0.0042$), and with lower surface areas of 7 out of 33 individual regions – superior parietal ($\beta = -0.0175$, $p_{FDR} = 0.0006$), precuneus ($\beta = -0.0131$, $p_{FDR} = 0.0089$), postcentral gyrus ($\beta = -0.0118$, $p_{FDR} = 0.0102$), fusiform ($\beta = -0.0103$, $p_{FDR} = 0.0296$), supramarginal ($\beta = -0.0099$, $p_{FDR} = 0.0296$), banks of superior temporal sulcus ($\beta = -0.0098$, $p_{FDR} = 0.0341$), and inferior parietal ($\beta = -0.0090$, $p_{FDR} = 0.0310$). No differences in cortical or subcortical volumes were found.

With regard to fiber-related white matter microstructure, late chronotype was associated with lower ICVF in association ($\beta = -0.0126$, $p_{FDR} = 0.0377$), projection ($\beta = -0.0118$, $p_{FDR} = 0.0377$), and thalamic fibers ($\beta = -0.0113$, $p_{FDR} = 0.0377$). Lower ICVF was also found in 5 out of 15 individual white matter tracts – superior thalamic radiation ($\beta = -0.0176$, $p_{FDR} = 0.0148$), uncinate fasciculus ($\beta = -0.0155$, $p_{FDR} = 0.0148$), corticospinal tract ($\beta = -0.0152$, $p_{FDR} = 0.0195$), cingulate gyrus ($\beta = -0.0152$, $p_{FDR} = 0.0148$), and superior longitudinal fasciculus ($\beta = -0.0150$, $p_{FDR} = 0.0195$). No differences in other measures of white matter microstructure were found.

###### S2.2.4 Brain Structure Differences Associated with *Ease of Getting Up*

At the global level, ease of getting up in the morning was associated with lower overall volume of white matter hyperintensities ($\beta = -0.0163$, $p = 0.0020$), and with higher global cortical surface area ($\beta = 0.0123$, $p < 0.0001$) and volume ($\beta = 0.0077$, $p = 0.0094$) measures.

At the lobar lever, ease of getting up was associated with lower occipital cortical thickness ($\beta = -0.0132$, $p_{FDR} = 0.0429$), but with larger frontal and parietal surface areas (respectively $\beta = 0.0121$, $p_{FDR} = 0.0002$ and $\beta = 0.0191$, $p_{FDR} < 0.0001$) and volumes (respectively $\beta = 0.0091$, $p_{FDR} = 0.0148$ and $\beta = 0.0143$, $p_{FDR} = 0.0002$). Higher surface area was also identified in the cingulate gyrus ($\beta = 0.0079$, $p_{FDR} = 0.0281$).

At the regional level, ease of getting up was associated with larger surface areas of 12 out of 33 individual regions, with biggest differences observed in superior parietal

($\beta = 0.0205$, $p_{FDR} < 0.0001$), postcentral ($\beta = 0.0170$, $p_{FDR} < 0.0001$), caudal middle frontal ($\beta = 0.0139$, $p_{FDR} = 0.0031$), supramarginal ($\beta = 0.0137$, $p_{FDR} = 0.0012$) and precuneus ($\beta = 0.0136$, $p_{FDR} = 0.0025$) regions. Higher cortical volumes were found in 7 of 33 regions – medial orbitofrontal ($\beta = 0.0141$, $p_{FDR} = 0.0040$), superior parietal ($\beta = 0.0134$, $p_{FDR} = 0.0143$), supramarginal ($\beta = 0.0122$, $p_{FDR} = 0.0108$), precuneus ($\beta = 0.0114$, $p_{FDR} = 0.0260$), postcentral ($\beta = 0.0110$, $p_{FDR} = 0.0343$), lateral orbitofrontal ($\beta = 0.0099$, $p_{FDR} = 0.0493$), and superior frontal ($\beta = 0.0094$, $p_{FDR} = 0.0447$).

With regard to subcortical structures, ease of getting up was associated with lower caudate and putamen volumes (respectively $\beta = -0.0207$, $p_{FDR} < 0.0001$ and $\beta = -0.0116$, $p_{FDR} = 0.0462$); no other associations were found.

With regard to white matter microstructure, ease of getting up was associated with higher OD of thalamic fibers ($\beta = 0.0161$, $p_{FDR} = 0.0077$) and higher OD in two individual tracts – corticospinal ($\beta = 0.0201$, $p_{FDR} = 0.0013$) and superior thalamic radiation ($\beta = 0.0195$, $p_{FDR} = 0.0013$).

##### *S2.3 Sensitivity Analysis Results*

Overall, sensitivity analysis results indicated that most differences in brain structure associated with *sleeping more than recommended, sleeping less than recommended*, or with *self-reported sleeplessness / insomnia* are independent of lifetime depression experience.

###### S2.3.1 Brain Structure Associations of Sleeping Longer than Recommended without Correction for Lifetime Depression

Without correction for lifetime depression, sleeping longer than recommended was no longer significantly associated with lower volumes of global white matter ( $p = 0.0596$ ), parahippocampal cortex ( $p = 0.1020$ ) and posterior cingulate cortex ( $p = 0.0736$ ). On the other hand, significant associations without correction were identified for lower global ICVF ( $\beta = -0.0124$, $p = 0.0443$ ), higher MD in projection fibers ( $\beta = 0.0118$, $p_{FDR} = 0.0489$ ), lower FA in forceps major ( $\beta = -0.0174$, $p_{FDR} = 0.0481$ ), and lower volume of the superior frontal cortex ( $\beta = -0.0116$, $p_{FDR} = 0.0169$ ). All other associations remained significant with similar effect magnitudes.

###### S2.3.2 Brain Structure Associations of Sleeping Less than Recommended without Correction for Lifetime Depression

After removing correction for lifetime depression, sleeping less than recommended was no longer significantly associated with lower cuneus and pericalcarine surface areas (respectively $p_{FDR} = 0.1014$ and $p_{FDR} = 0.0519$ ), lower volume of the rostral anterior cingulate ( $p_{FDR} = 0.0610$ ), or lower MD in middle cerebellar peduncle ( $p_{FDR} = 0.0622$ ). All other associations remained significant.

###### S2.3.3 Brain Structure Associations of Insomnia/Sleeplessness without Correction for Lifetime Depression

All significant brain structure association of self-reported insomnia/sleeplessness remained significant after removing correction for lifetime depression status, with similar effect magnitudes.

###### S2.3.4 Brain Structure Associations of Sleeping Longer than Recommended without Correction for Health Conditions

Without correction for all health-related covariates (please see section S1.4 above), sleeping longer than recommended was no longer significantly associated with lower overall white matter volume ($p = 0.1654$), but was instead significantly associated with lower global ICVF ($\beta = -0.0130,\ p = 0.0321$), lower surface area of the occipital lobe ($\beta = -0.0124,\ p_{FDR} = 0.0354$), lower FA in association and thalamic fibers (respectively $\beta = -0.0137,\ p_{FDR} = 0.0372$ and $\beta = -0.0138,\ p_{FDR} = 0.0372$), lower ICVF in association fibers ($\beta = -0.0131,\ p_{FDR} = 0.0463$), and also higher MD and ISOVF in projection fibers (respectively $\beta = 0.0172,\ p_{FDR} = 0.0036$ and $\beta = 0.0176,\ p_{FDR} = 0.0058$).

At the level of individual regions, additional significant associations without correction were identified for thickness of the posterior cingulate cortex ($\beta = -0.0117,\ p_{FDR} = 0.0236$), volume of the ventral diencephalon ($\beta = -0.0125,\ p_{FDR} = 0.0089$), and volumes of 8 additional cortical regions (largest effects – superior frontal $\beta = -0.0130,\ p_{FDR} = 0.0035$, lingual $\beta = -0.0127,\ p_{FDR} = 0.0263$, postcentral $\beta = -0.0109,\ p_{FDR} = 0.0263$).

At the level of individual white matter tracts, additional significant associations were found for FA in forceps major ($\beta = -0.0169,\ p_{FDR} = 0.0403$), forceps minor ($\beta = -0.0149,\ p_{FDR} = 0.0403$), inferior frontooccipital fasciculus ($\beta = -0.0141,\ p_{FDR} = 0.0403$), and corticospinal tract ($\beta = 0.0141,\ p_{FDR} = 0.0403$). Additional higher MD was found in middle cerebellar peduncle ($\beta = 0.0172,\ p_{FDR} = 0.0098$) and superior longitudinal fasciculus ($\beta = 0.0154,\ p_{FDR} = 0.0108$).

###### S2.3.5 Brain Structure Associations of Sleeping Less than Recommended without

**Correction for Health Conditions**

Without correction for health-related covariates, all previously identified associations of global brain measures with sleeping less than recommended remained significant. Additional significant associations were revealed for surface areas of temporal and parietal lobes ($\beta = -0.0065,\ p_{FDR} = 0.0246$ and $\beta = -0.0072,\ p_{FDR} = 0.0246$), volumes of frontal and temporal lobes ($\beta = -0.0072,\ p_{FDR} = 0.0343$ and $\beta = -0.0063,\ p_{FDR} = 0.0463$), FA of thalamic fibers ($\beta = -0.0115,\ p_{FDR} = 0.0470$), and OD of association fibers ($\beta = 0.0122,\ p_{FDR} = 0.0250$).

At the level of individual regions, additional significant associations without correction were identified for surface areas of fusiform ($\beta = -0.0092,\ p_{FDR} = 0.0193$), middle temporal ($\beta = -0.0091,\ p_{FDR} = 0.0159$), inferior temporal ($\beta = -0.0089,\ p_{FDR} = 0.0264$), inferior parietal ($\beta = -0.0085,\ p_{FDR} = 0.0183$) and pars orbitalis ($\beta = -0.0081,\ p_{FDR} = 0.0183$). Additional significant associations were also found for volumes or pericalcarine ($\beta = -0.0134,\ p_{FDR} = 0.0175$), caudal middle frontal ($\beta = -0.0113,\ p_{FDR} = 0.0175$), lateral orbitofrontal ($\beta = -0.0107,\ p_{FDR} = 0.0174$), middle temporal ($\beta = -0.0105,\ p_{FDR} = 0.0174$), and rostral middle frontal ($\beta = -0.0093,\ p_{FDR} = 0.0315$) cortical regions.

With regard to subcortical region volumes, association with cerebellar white matter volume was no longer significant without correction for health-related covariates ($p_{FDR} = 0.0905$), but a significant association with volume of the putamen was identified ($\beta = 0.0121,\ p_{FDR} = 0.0391$).

At the level of individual white matter tracts, association with MD of the middle cerebellar peduncle was no longer significant without correction ( $p_{FDR} = 0.0321$ ), but an additional significant association was found for lower FA in superior longitudinal fasciculus ( $\beta = -0.0121$, $p_{FDR} = 0.0291$ ).

###### S2.3.6 Brain Structure Associations of Insomnia/Sleeplessness without Correction for Health Conditions

Without correction for health-related covariates, association of insomnia/sleeplessness with overall grey matter volume was no longer significant ( $p_{FDR} = 0.0596$ ), but additional significant associations were found for lower frontal cortical lobar volume ( $\beta = -0.0126$, $p_{FDR} = 0.0180$ ), lower lateral orbitofrontal cortical thickness ( $\beta = -0.0225$, $p_{FDR} = 0.0136$ ), and lower volume of the precentral cortex ( $\beta = -0.0179$, $p_{FDR} = 0.0189$ ).

**Table S1**

**Assignment of Desikan-Killiany cortical regions to five lobes**

| Cortical Region | Frontal Lobe | Temporal Lobe | Parietal Lobe | Cingulate Lobe | Occipital Lobe |
|---|---|---|---|---|---|
| Caudal Middle Frontal | √ | - | - | - | - |
| Lateral Orbitofrontal | √ | - | - | - | - |
| Medial Orbitofrontal | √ | - | - | - | - |
| Paracentral | √ | - | √ | - | - |
| Pars Opercularis | √ | - | - | - | - |
| Pars Orbitalis | √ | - | - | - | - |
| Pars Triangularis | √ | - | - | - | - |
| Precentral | √ | - | - | - | - |
| Rostral Middle Frontal | √ | - | - | - | - |
| Superior Frontal | √ | - | - | - | - |
| Frontal Pole | √ | - | - | - | - |
| Banks of Superior Temporal Sulcus | - | √ | - | - | - |
| Entorhinal | - | √ | - | - | - |
| Fusiform | - | √ | - | - | - |
| Inferior Temporal | - | √ | - | - | - |
| Middle Temporal | - | √ | - | - | - |
| Parahippocampal | - | √ | - | - | - |
| Superior Temporal | - | √ | - | - | - |
| Transverse Temporal | - | √ | - | - | - |
| Insula | - | √ | - | - | - |
| Inferior Parietal | - | - | √ | - | - |
| Postcentral | - | - | √ | - | - |
| Precuneus | - | - | √ | - | - |
| Superior Parietal | - | - | √ | - | - |
| Supramarginal | - | - | √ | - | - |
| Caudal Anterior Cingulate | - | - | - | √ | - |
| Rostral Anterior Cingulate | - | - | - | √ | - |
| Isthmus Cingulate | - | - | - | √ | - |
| Posterior Cingulate | - | - | - | √ | - |
| Cuneus | - | - | - | - | √ |
| Lateral Occipital | - | - | - | - | √ |
| Lingual | - | - | - | - | √ |
| Pericalcarine | - | - | - | - | √ |

***Note***: *Paracentral* cortex is the only region assigned to more than one lobe (frontal and parietal lobes).

**Table S2**

**Assignment of individual white matter tracts to white matter fiber bundles**

| White Matter Tract | Global White Matter | Association Fibers | Projection Fibers | Thalamic Fibers |
|---|---|---|---|---|
| Cingulate Gyrus Left | √ | √ | - | - |
| Cingulate Gyrus Right | √ | √ | - | - |
| Forceps Major | √ | √ | - | - |
| Forceps Minor | √ | √ | - | - |
| Inferior Frontooccipital Fasciculus Left | √ | √ | - | - |
| Inferior Frontooccipital Fasciculus Right | √ | √ | - | - |
| Inferior Longitudinal Fasciculus Left | √ | √ | - | - |
| Inferior Longitudinal Fasciculus Right | √ | √ | - | - |
| Superior Longitudinal Fasciculus Left | √ | √ | - | - |
| Superior Longitudinal Fasciculus Right | √ | √ | - | - |
| Parahippocampal Left | √ | √ | - | - |
| Parahippocampal Right | √ | √ | - | - |
| Uncinate Fasciculus Left | √ | √ | - | - |
| Uncinate Fasciculus Right | √ | √ | - | - |
| Acoustic Radiation Left | √ | - | √ | - |
| Acoustic Radiation Right | √ | - | √ | - |
| Corticospinal Tract Left | √ | - | √ | - |
| Corticospinal Tract Right | √ | - | √ | - |
| Medial Lemniscus Left | √ | - | √ | - |
| Medial Lemniscus Right | √ | - | √ | - |
| Middle Cerebellar Peduncle | √ | - | √ | - |
| Anterior Thalamic Radiation Left | √ | - | - | √ |
| Anterior Thalamic Radiation Right | √ | - | - | √ |
| Posterior Thalamic Radiation Left | √ | - | - | √ |
| Posterior Thalamic Radiation Right | √ | - | - | √ |
| Superior Thalamic Radiation Left | √ | - | - | √ |
| Superior Thalamic Radiation Right | √ | - | - | √ |

**Table S3**

**Loadings (coefficients) of individual white matter tract FA measures onto global and fiber-related FA measures derived with PCA**

| FA Measure Tract | gFA Global Loading | gFA Association Loading | gFA Projection Loading | gFA Thalamic Loading |
|---|---|---|---|---|
| Cingulate Gyrus Left | 0.26022 | 0.36154 | - | - |
| Cingulate Gyrus Right | 0.24403 | 0.34189 | - | - |
| Forceps Major | 0.20992 | 0.25968 | - | - |
| Forceps Minor | 0.21717 | 0.27279 | - | - |
| Inferior Frontooccipital Fasciculus Left | 0.24326 | 0.30329 | - | - |
| Inferior Frontooccipital Fasciculus Right | 0.23176 | 0.28571 | - | - |
| Inferior Longitudinal Fasciculus Left | 0.21307 | 0.26639 | - | - |
| Inferior Longitudinal Fasciculus Right | 0.19535 | 0.24326 | - | - |
| Superior Longitudinal Fasciculus Left | 0.18910 | 0.23817 | - | - |
| Superior Longitudinal Fasciculus Right | 0.18538 | 0.23175 | - | - |
| Parahippocampal Left | 0.15719 | 0.20897 | - | - |
| Parahippocampal Right | 0.15094 | 0.19973 | - | - |
| Uncinate Fasciculus Left | 0.19278 | 0.24833 | - | - |
| Uncinate Fasciculus Right | 0.17480 | 0.22615 | - | - |
| Acoustic Radiation Left | 0.18548 | - | 0.3388 | - |
| Acoustic Radiation Right | 0.17857 | - | 0.33559 | - |
| Corticospinal Tract Left | 0.20653 | - | 0.47375 | - |
| Corticospinal Tract Right | 0.20882 | - | 0.48456 | - |
| Medial Lemniscus Left | 0.079351 | - | 0.25566 | - |
| Medial Lemniscus Right | 0.090112 | - | 0.27577 | - |
| Middle Cerebellar Peduncle | 0.16598 | - | 0.41466 | - |
| Anterior Thalamic Radiation Left | 0.19418 | - | - | 0.41173 |
| Anterior Thalamic Radiation Right | 0.18879 | - | - | 0.40131 |
| Posterior Thalamic Radiation Left | 0.20391 | - | - | 0.4681 |
| Posterior Thalamic Radiation Right | 0.2039 | - | - | 0.46038 |
| Superior Thalamic Radiation Left | 0.15017 | - | - | 0.33155 |
| Superior Thalamic Radiation Right | 0.16172 | - | - | 0.35838 |

**Table S4**

**Loadings (coefficients) of individual white matter tract MD measures onto global and fiber-related MD measures derived with PCA**

| MD Measure Tract | gMD Global Loading | gMD Association Loading | gMD Projection Loading | gMD Thalamic Loading |
|---|---|---|---|---|
| Cingulate Gyrus Left | 0.14822 | 0.18359 | - | - |
| Cingulate Gyrus Right | 0.14645 | 0.17899 | - | - |
| Forceps Major | 0.20785 | 0.27111 | - | - |
| Forceps Minor | 0.19329 | 0.22809 | - | - |
| Inferior Frontooccipital Fasciculus Left | 0.22686 | 0.27352 | - | - |
| Inferior Frontooccipital Fasciculus Right | 0.22928 | 0.28058 | - | - |
| Inferior Longitudinal Fasciculus Left | 0.21929 | 0.27064 | - | - |
| Inferior Longitudinal Fasciculus Right | 0.21052 | 0.26138 | - | - |
| Superior Longitudinal Fasciculus Left | 0.18451 | 0.22238 | - | - |
| Superior Longitudinal Fasciculus Right | 0.18066 | 0.22013 | - | - |
| Parahippocampal Left | 0.24527 | 0.38902 | - | - |
| Parahippocampal Right | 0.24632 | 0.39304 | - | - |
| Uncinate Fasciculus Left | 0.17719 | 0.24279 | - | - |
| Uncinate Fasciculus Right | 0.16898 | 0.22573 | - | - |
| Acoustic Radiation Left | 0.15425 | - | 0.29254 | - |
| Acoustic Radiation Right | 0.16462 | - | 0.28126 | - |
| Corticospinal Tract Left | 0.14185 | - | 0.22895 | - |
| Corticospinal Tract Right | 0.14224 | - | 0.23428 | - |
| Medial Lemniscus Left | 0.082403 | - | 0.30413 | - |
| Medial Lemniscus Right | 0.079879 | - | 0.32383 | - |
| Middle Cerebellar Peduncle | 0.18298 | - | 0.72845 | - |
| Anterior Thalamic Radiation Left | 0.2349 | - | - | 0.41715 |
| Anterior Thalamic Radiation Right | 0.2322 | - | - | 0.41273 |
| Posterior Thalamic Radiation Left | 0.25799 | - | - | 0.49574 |
| Posterior Thalamic Radiation Right | 0.25587 | - | - | 0.47773 |
| Superior Thalamic Radiation Left | 0.16083 | - | - | 0.29632 |
| Superior Thalamic Radiation Right | 0.16501 | - | - | 0.30634 |

**Table S5**

**Loadings (coefficients) of individual white matter tract ICVF measures onto global and fiber-related ICVF measures derived with PCA**

| ICVF Measure Tract | gICVF Global Loading | gICVF Association Loading | gICVF Projection Loading | gICVF Thalamic Loading |
|---|---|---|---|---|
| Cingulate Gyrus Left | 0.21558 | 0.27971 | - | - |
| Cingulate Gyrus Right | 0.20143 | 0.2603 | - | - |
| Forceps Major | 0.19786 | 0.249 | - | - |
| Forceps Minor | 0.23882 | 0.3116 | - | - |
| Inferior Frontooccipital Fasciculus Left | 0.23892 | 0.30678 | - | - |
| Inferior Frontooccipital Fasciculus Right | 0.23979 | 0.30802 | - | - |
| Inferior Longitudinal Fasciculus Left | 0.22288 | 0.28848 | - | - |
| Inferior Longitudinal Fasciculus Right | 0.21564 | 0.28065 | - | - |
| Superior Longitudinal Fasciculus Left | 0.22937 | 0.30111 | - | - |
| Superior Longitudinal Fasciculus Right | 0.22431 | 0.29489 | - | - |
| Parahippocampal Left | 0.13648 | 0.1751 | - | - |
| Parahippocampal Right | 0.13504 | 0.17297 | - | - |
| Uncinate Fasciculus Left | 0.17647 | 0.23291 | - | - |
| Uncinate Fasciculus Right | 0.17159 | 0.22691 | - | - |
| Acoustic Radiation Left | 0.19371 | - | 0.42807 | - |
| Acoustic Radiation Right | 0.19043 | - | 0.42721 | - |
| Corticospinal Tract Left | 0.16168 | - | 0.40224 | - |
| Corticospinal Tract Right | 0.1621 | - | 0.40532 | - |
| Medial Lemniscus Left | 0.083059 | - | 0.22974 | - |
| Medial Lemniscus Right | 0.085848 | - | 0.24699 | - |
| Middle Cerebellar Peduncle | 0.13505 | - | 0.44089 | - |
| Anterior Thalamic Radiation Left | 0.21359 | - | - | 0.42796 |
| Anterior Thalamic Radiation Right | 0.20985 | - | - | 0.4228 |
| Posterior Thalamic Radiation Left | 0.21085 | - | - | 0.42309 |
| Posterior Thalamic Radiation Right | 0.21606 | - | - | 0.43225 |
| Superior Thalamic Radiation Left | 0.18 | - | - | 0.3629 |
| Superior Thalamic Radiation Right | 0.18363 | - | - | 0.3749 |

**Table S6**

**Loadings (coefficients) of individual white matter tract OD measures onto global and fiber-related OD measures derived with PCA**

| OD Measure Tract | gOD Global Loading | gOD Association Loading | gOD Projection Loading | gOD Thalamic Loading |
|---|---|---|---|---|
| Cingulate Gyrus Left | 0.11597 | 0.11481 | - | - |
| Cingulate Gyrus Right | 0.13471 | 0.13875 | - | - |
| Forceps Major | 0.073441 | 0.062245 | - | - |
| Forceps Minor | 0.10527 | 0.084268 | - | - |
| Inferior Frontooccipital Fasciculus Left | 0.13946 | 0.13515 | - | - |
| Inferior Frontooccipital Fasciculus Right | 0.14357 | 0.13441 | - | - |
| Inferior Longitudinal Fasciculus Left | 0.12085 | 0.11925 | - | - |
| Inferior Longitudinal Fasciculus Right | 0.11842 | 0.11885 | - | - |
| Superior Longitudinal Fasciculus Left | 0.10599 | 0.06241 | - | - |
| Superior Longitudinal Fasciculus Right | 0.117 | 0.072069 | - | - |
| Parahippocampal Left | 0.45143 | 0.61442 | - | - |
| Parahippocampal Right | 0.49729 | 0.64967 | - | - |
| Uncinate Fasciculus Left | 0.1915 | 0.20774 | - | - |
| Uncinate Fasciculus Right | 0.18389 | 0.20022 | - | - |
| Acoustic Radiation Left | 0.15679 | - | 0.27632 | - |
| Acoustic Radiation Right | 0.17814 | - | 0.27116 | - |
| Corticospinal Tract Left | 0.15413 | - | 0.31985 | - |
| Corticospinal Tract Right | 0.16854 | - | 0.3312 | - |
| Medial Lemniscus Left | 0.060443 | - | 0.080855 | - |
| Medial Lemniscus Right | 0.088457 | - | 0.10482 | - |
| Middle Cerebellar Peduncle | 0.22658 | - | 0.78778 | - |
| Anterior Thalamic Radiation Left | 0.16681 | - | - | 0.2839 |
| Anterior Thalamic Radiation Right | 0.18367 | - | - | 0.3583 |
| Posterior Thalamic Radiation Left | 0.12428 | - | - | 0.14078 |
| Posterior Thalamic Radiation Right | 0.13143 | - | - | 0.16725 |
| Superior Thalamic Radiation Left | 0.18047 | - | - | 0.59424 |
| Superior Thalamic Radiation Right | 0.19648 | - | - | 0.62459 |

**Table S7**

**Loadings (coefficients) of individual white matter tract ISOVF measures onto global and fiber-related ISOVF measures derived with PCA**

| ISOVF Measure Tract | gISOVF Global Loading | gISOVF Association Loading | gISOVF Projection Loading | gISOVF Thalamic Loading |
|---|---|---|---|---|
| Cingulate Gyrus Left | 0.12011 | 0.10436 | - | - |
| Cingulate Gyrus Right | 0.10257 | 0.085575 | - | - |
| Forceps Major | 0.14453 | 0.11828 | - | - |
| Forceps Minor | 0.11356 | 0.074151 | - | - |
| Inferior Frontooccipital Fasciculus Left | 0.14036 | 0.099326 | - | - |
| Inferior Frontooccipital Fasciculus Right | 0.15711 | 0.11687 | - | - |
| Inferior Longitudinal Fasciculus Left | 0.14161 | 0.097766 | - | - |
| Inferior Longitudinal Fasciculus Right | 0.16219 | 0.11804 | - | - |
| Superior Longitudinal Fasciculus Left | 0.13749 | 0.088955 | - | - |
| Superior Longitudinal Fasciculus Right | 0.16113 | 0.10937 | - | - |
| Parahippocampal Left | 0.50327 | 0.65743 | - | - |
| Parahippocampal Right | 0.49592 | 0.65922 | - | - |
| Uncinate Fasciculus Left | 0.12838 | 0.12257 | - | - |
| Uncinate Fasciculus Right | 0.12507 | 0.11657 | - | - |
| Acoustic Radiation Left | 0.072695 | - | 0.0629 | - |
| Acoustic Radiation Right | 0.11047 | - | 0.052215 | - |
| Corticospinal Tract Left | 0.098139 | - | 0.072913 | - |
| Corticospinal Tract Right | 0.11444 | - | 0.09008 | - |
| Medial Lemniscus Left | 0.07346 | - | 0.34204 | - |
| Medial Lemniscus Right | 0.063986 | - | 0.34914 | - |
| Middle Cerebellar Peduncle | 0.22862 | - | 0.86081 | - |
| Anterior Thalamic Radiation Left | 0.17844 | - | - | 0.31311 |
| Anterior Thalamic Radiation Right | 0.16762 | - | - | 0.29491 |
| Posterior Thalamic Radiation Left | 0.19667 | - | - | 0.58644 |
| Posterior Thalamic Radiation Right | 0.19772 | - | - | 0.54592 |
| Superior Thalamic Radiation Left | 0.1271 | - | - | 0.28927 |
| Superior Thalamic Radiation Right | 0.1359 | - | - | 0.29896 |
